# Potential Misrepresentation of Inherited Breast Cancer Risk by Common Germline Alleles

**DOI:** 10.1101/2022.10.21.22281361

**Authors:** William Letsou, Fan Wang, Wonjong Moon, Cindy Im, Yadav Sapkota, Leslie L. Robison, Yutaka Yasui

**Affiliations:** Department of Epidemiology and Cancer Control, St. Jude Children’s Research Hospital, Memphis, TN 38105, USA; School of Public Health, University of Alberta, Edmonton, AB T6G 1C9, Canada

**Keywords:** Haplotypes, pattern mining, breast cancer, common variant, UK Biobank

## Abstract

Hundreds of common variants have been found to confer small but significant differences in breast cancer risk, supporting the polygenic additive model of inherited risk. This widely accepted model is at odds with twin data indicating highly elevated risk in a subgroup of women. Using a novel closed-pattern-mining algorithm, we provide evidence that rare variants or haplotypes may underlie the association of breast cancer risk with common germline alleles. Our method, called Chromosome Overlap, consists in iteratively pairing chromosomes from affected individuals and looking for noncontiguous patterns of shared alleles without exhaustive enumeration. We applied Chromosome Overlap to haplotypes of genotyped SNPs from 9,011 female breast cancer cases from the UK Biobank (UKBB) at three topologically associating domains containing well-established common-allele “hits” for breast cancer. A total of 181,034 UKBB women of “white British” ancestry were used to assess the discovered haplotypes, and 55,346 cases and controls of European ancestry in the Discovery, Biology, and Risk of Inherited Variants in Breast Cancer (DRIVE) case-control study were used for replication. Out of twenty rare (frequency < ∼0.1%) risk haplotypes of large effect identified in UKBB at *P* < 1.0 × 10^−5^, four (hazard ratio: 4.22–20.2) were subsequently replicated in DRIVE (odds ratio: 2.13–11.9) at *P* < 0.05. Our results support the genetic heterogeneity and rare-variant/haplotype basis of breast cancer risk and suggest a novel type of “synthetic association” wherein common risk alleles on a rare risk haplotype may misrepresent disease risk through their tagging of many “false positive” haplotypes.

**Significance:** Chromosome Overlap reveals that common alleles identified by GWAS may be poor surrogates for underlying high-risk haplotypes, necessitating a reappraisal of the polygenic model of disease risk.

## Introduction

Genome-wide association studies (GWAS) have discovered many single nucleotide polymorphisms (SNPs) associated with small, yet robustly replicated, differences in breast cancer risk (1,2). Polygenic risk scores (PRSs) of these common variants can differentiate women’s breast cancer risk by up to several fold (3). While the PRS is useful for risk stratification in a population, the causal germline genetic variation underlying many of these GWAS hits remains unclear. For example, while SNPs on DNA microarrays are designed to tag common haplotypes, it is not clear whether the haplotypes tagged by GWAS hits are causal or if there are haplotypes or rare variants of large effect correlated, perhaps weakly, with the GWAS hits (hence their small effect sizes) that truly cause changes in breast cancer risk. Here we consider the latter hypothesis that rare haplotypes or variants of large effect underlie the associations of the GWAS hits with breast cancer risk. This hypothesis addresses two key inconsistencies of the prevailing additive, polygenic, common-variant model for breast cancer risk (1-4) that is seemingly supported by the evidence of GWAS hits. One is the remarkably high incidence rates in first-degree relatives of breast cancer cases (5). This observation is incompatible with common SNPs being the determinants of breast cancer risk but is consistent with rare haplotypes or mutations of large effect (6). The other is the equality of monozygotic and dizygotic twins’ breast cancer incidence interval when both twins develop breast cancer (7). This observation suggests that, when both twins develop breast cancer, there is no difference between monozygotic and dizygotic twins regarding their overall genetic susceptibility, a conclusion which is incompatible with the polygenic model of inheritance but consistent with the hypothesis that the specific genetic risk factor in a given family is *monogenic*, coming from a single locus (6).

Related hypotheses have been proposed attempting to explain similar shortcomings in the polygenic model. One is the hypothesis that so-called “synthetic associations” of rare variants near GWAS hits explain observed GWAS signals (8). While theoretical and sequencing studies have not provided strong evidence for this hypothesis (9-12), it has not been considered whether rare haplotypes, instead of rare variants, underlie GWAS hits. Another hypothesis is that human diseases have substantial “genetic heterogeneity,” being due primarily to rare mutations, such as those of the *BRCA1*/*2* genes, which segregate in families (13). Neither of these hypotheses explicitly consider the possibility that rare haplotypes composed of SNPs may be tags for rare causal variants or haplotypes, or, by virtue of their particular allele combinations, causal themselves.

The problem of looking for risk haplotypes is combinatorial complexity: there will be a total of 3^*m*^ − 1 possible non-contiguous haplotypes for every *m* independent SNPs. We avoid this combinatorial explosion using tools from the pattern-mining literature (14-16). Our approach, called Chromosome Overlap, is to iteratively pair chromosomes from affected individuals and look for shared sets of alleles. The resulting patterns, called *closed patterns*, are then tested for association with breast cancer risk. Closed-pattern mining and related techniques that avoid the task of exhaustive enumeration have been used previously to detect genetic epistasis (17-19), but not to discover risk haplotypes composed of SNPs underlying GWAS hits. We applied Chromosome Overlap to phased genotype data in the UK Biobank (UKBB) (20) to discover risk haplotypes of large effect composed of SNPs in the vicinity of three of the strongest GWAS hits (by *p*-value) in the GWAS Catalog (https://www.ebi.ac.uk/gwas/home) associated with breast cancer risk: rs2981578 on chromosome 10q26 in an intron of *FGFR2* (21); rs554219 on chromosome 11q13 upstream of *CCND1* (22); and rs4784227 on 16q12 in an intron of *CASC16* (23). Subsequently, we replicated several of these haplotypes in the Discovery, Biology, and Risk of Inherited Variants in Breast Cancer (DRIVE) case-control study (http://epi.grants.cancer.gov/gameon/), generating support for the genetic heterogeneity of breast cancer and the notion of synthetic associations of rare haplotypes with GWAS hits.

## Materials and Methods

### Chromosome Overlap: An iterative, parallel procedure for discovering risk haplotypes

For this paper, a *pattern* is a subset of (not necessarily contiguous) SNP-alleles, and a chromosome is said to contain a pattern if each of its alleles agrees with the alleles of the pattern. A pattern is called *closed* if it is the longest pattern shared by a set of chromosomes (15). For example, Abd is the longest pattern shared by chromosomes *AbCde, AbCdE* and *AbcdE*. Closed patterns were introduced by Pasquier *et al*. (14) as a minimal but lossless representation of the frequent transactions in a database. Terada and colleagues used a linear-time closed-pattern-mining algorithm, called LCM (16), in conjunction with a multiple-testing procedure to find gene-expression and SNP patterns that are associated with different phenotypes (18,19,45). A limitation of their approach is that it only identifies patterns consisting of minor alleles (45) and builds patterns from the bottom up (i.e., short to long).

Recognizing that risk haplotypes may be very long and comprise reference and alternate alleles alike, we developed a new pattern-mining method, called Chromosome Overlap, to select and test haplotypes of common SNP-alleles in phased genotype data for breast cancer risk association. Our method is a top-down scheme that forms combinations of chromosomes from affected individuals and looks for shared alleles. The shared alleles of a σ-tuple of chromosomes constitute a closed pattern called a *meta-chromosome* (Fig. 1A). Meta-chromosomes can themselves be overlapped to find additional closed patterns (Fig. 1C). The following steps, described with additional mathematical details in the Supplementary Methods Section 1–3, are used to discover the complete set of closed patterns:

**Figure 1:**
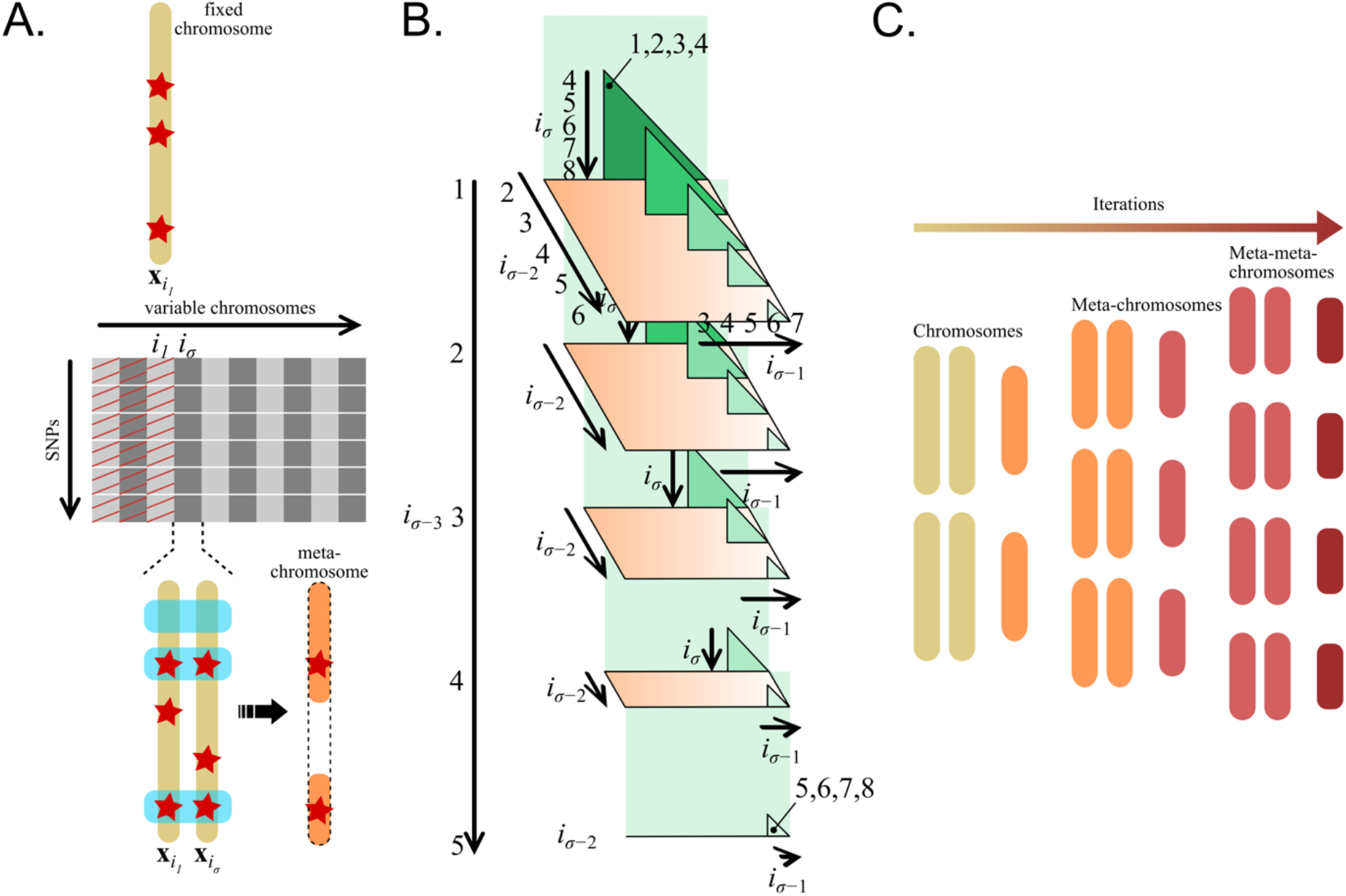
The Chromosome Overlap method. (A) Illustration of the initiation step forming meta-chromosomes in the case *σ* = 2. One chromosome (**x**_*i*1_ = **x**_3_) is fixed and overlapped with all variable chromosomes **x**_*j*_ satisfying *i*_*σ*_ = *j* > 3 in the non-hatched region of the haplotype matrix. The operation in which **x**_3_ forms a non-contiguous meta-chromosome with column *i*_2_ = 4 is shown in detail. (B) A general triangular array for choosing σ-tuples of chromosomes from a fixed total, which can be used to partition of all possible overlap operations to different computing nodes. The figure shows how all combinations of *σ* = 4 chromosomes (e.g., (1,2,3,4), (1,2,5,8), (5,6,7,8)) from a fixed total of 8 can be indexed. The lowest index *i*_*σ* – 3_ = *i*_1_ takes all values from 1 to 8 − *σ* + (*σ* − 3) = 5, corresponding to the layers of the array, and in general, index *i*_*σ* − *j*_ spans the integers *i*_*σ* − *j* − 1_ + 1 to 8 − *j* in the *j* + 1^th^ dimension. The process may be generalized to any value of *σ* by adding the appropriate number of dimensions. (C) A schematic representation of the overlap iterations with *σ* = 2 (pairing), illustrating the overlap of chromosome pairs to form meta-chromosomes and the overlap of meta-chromosome pairs to form meta-meta-chromosomes. Note that, in general, meta-chromosomes need be neither contiguous nor of the same length.

1. Initiation phase forming all pairs (*σ* = 2) of chromosomes of breast cancer cases and identifying shared allele patterns in a genomic region of interest.
2. Filtering of the shared patterns (meta-chromosomes) that pass a statistical-significance cutoff in the association analysis of breast cancer risk determined to prevent a combinatorial explosion.
3. Iterated pairwise overlap of meta-chromosomes until all closed patterns (among the filtered set from step 2) have been found.

A key feature of this method, which distinguishes it from earlier top-down approaches (15), is a triangular-array algorithm (Fig. 1B; Supplementary Methods Section 2) that allows the overlap operations to be parallelized among different computing nodes without exhaustive enumeration of a combinatorically large set. Chromosome Overlap was designed to run on IBM Spectrum LSF for distributed high-performance computing environments, but sample code executable on a single machine is available at https://github.com/wletsou/ChromosomeOverlap.

### Haplotype data

Phased haplotype data from the UK Biobank (UKBB) was used for the discovery analysis. As described previously (20), this phased dataset consists of 487,409 samples phased at 658,720 autosomal SNPs on the Applied Biosystems UK BiLEVE Axiom Array. A total of 181,034 women who were designated as “white British” in UKBB were included in our analysis after excluding those who: (1) were identified to be outliers in heterozygosity or genotype missingness rates; (2) showed any sex chromosome aneuploidies; (3) were second-degree or closer relatives of any or third-degree relatives of more than ten other genotyped individuals; or (4) withdrew from UKBB before this study began. Genotype principal components for the discovery analysis in the 181,034 women were provided by the UK Biobank from a larger set of 407,219 individuals at 147,604 genotyped markers (Supplementary Fig. S2). The preceding steps were performed in PLINK (46) and KING (47). Breast cancer (UKBB data-field 40006, ICD10 code C50) was reported in 9,011 women with a mean (SD) age of onset of 56.2 (8.6) years; the remaining 172,023 women free of breast cancer had mean (SD) age of 65.0 (7.9) years at the end of follow-up.

Female subjects in the Discovery, Biology, and Risk of Inherited Variants in Breast Cancer (DRIVE) study were used for replication. Briefly, the DRIVE study was initiated in 2010 as part of the NCI’s Genetic Associations and Mechanisms in Oncology initiative (http://epi.grants.cancer.gov/gameon/) and includes data from 60,015 breast cancer cases and controls genotyped on the custom Illumina OncoArray (48). The 528,620 OncoArray SNPs were filtered to remove SNPs that: (1) had genotype missingness rate >10%; (2) were monomorphic; or (3) were not in Hardy-Weinberg equilibrium (*P* < 1 × 10^−10^), leaving a total of 433,297. A total of 4,669 subjects were excluded who: (1) showed any sex chromosome aneuploidies; (2) had genotyping rate <90%; (3) were identified as male by PLINK’s sex check; (4) were second-degree or closer relatives of any other subject; (5) separated from the European cluster in PCA (Supplementary Fig. S3); or (6) had missing age data. The preceding steps were performed in PLINK (46), KING (47), and FlashPCA2 (49), leaving 30,064 cases (mean (SD) age at onset: 61.7 (10.7) years) and 25,282 controls (mean (SD) age at end of follow-up: 59.6 (10.7) years).

Because SNPs were genotyped on different arrays in UKBB and DRIVE, we carried out genotype imputation to improve the coverage of UKBB Axiom SNPs on the DRIVE OncoArray. Imputation was performed in three batches of 15,000–20,000 samples using the TOPMed Imputation Server (https://imputation.biodatacatalyst.nhlbi.nih.gov) running Minimac4 (50) and phased using Eagle2 (51), based on the TOPMed reference panel including 194,512 haplotypes and 308 million sequenced variants (https://topmed.nhlbi.nih.gov) (52). In order to improve phasing consistency between UKBB and DRIVE, UKBB subjects were phased and imputed through the TOPMed pipeline; UKBB-phased genotypes were not used. PCA based on the imputed markers was used to generate genotype principal components for the replication analysis (Supplementary Figs. S4–S5). However, only the subset of imputed SNPs that were (1) genotyped on the UKBB Axiom Array, (2) imputed with Minimac’s quality statistic Rsq ≥ 0.8, and (3) biallelic in both cohorts were used to form haplotypes in the vicinity of the three strong GWAS hits.

### Statistical analysis

For the UKBB discovery analysis, we used Cox proportional-hazards models, adjusted for the first ten genotype principal components, with age as the time axis to assess the association of the discovered pattern(s) (either a single haplotype or multiple) with breast cancer incidence, where the number (zero, one, or two) of copies of each candidate haplotype was modeled as a continuous variable, as per the additive genetic model. The adjusted hazard ratio (HR) and *P*-value per haplotype copy were estimated using the one-degree-of-freedom likelihood ratio test (LRT) compared to the model without the haplotype. Because there were many overlapping, correlated patterns, we used stepwise forward-selection to add candidate haplotypes sequentially that had the smallest *P*-value until no candidate haplotype had *P* < 1 × 10^−5^. The reported p-values of the haplotypes were found by removing each surviving haplotype individually and computing the LRT statistic.

To reduce the number of SNP-alleles that defined each haplotype without changing the carrier/non-carrier status of each UKBB woman (i.e., to remove SNP-alleles unnecessary for indicating the same haplotype carriers), we used rpart (24), the recursive partitioning procedure implemented in R, with parameters cp = −1 and minsplit = 1 to ensure that the full tree was grown within the 30 steps allowed by R. SNP-alleles for splitting at each step were chosen to maximize the Gini impurity reduction of the node. Reduction was deemed necessary for removing extraneous SNP-alleles from common haplotypes in Phase 1 but was not used for rare haplotypes in Phase 2 where frequencies in the replication population could be sensitive to the removal of even a few SNP-alleles.

For the DRIVE replication analysis, we used logistic regression, adjusting for age and the first ten genotype principal components, and Fisher’s Exact test to assess the patterns (i.e., haplotypes) identified in the discovery analysis for association with breast cancer risk (DRIVE is not a cohort study, so Cox regression was inappropriate). The odds ratio (OR) and *P*-value per haplotype copy were estimated using the one-degree-of-freedom LRT (compared to the model without the haplotype) for each haplotype individually or using Fisher’s Exact test when the expected counts fell below 5 in any cell of the 2 × 2 table, with *P* < 0.05 deemed a successful replication. In the event that controls haplotype frequencies were not statistically significantly different between DRIVE and UKBB (Fisher Exact Test *P* ≥ 0.05), a combined analysis was carried out in a similar manner using DRIVE cases versus DRIVE and UKBB controls, adjusting for age and ten genotype principal components of the combined cohort for sufficiently common haplotype or using Fisher’s Exact test when the expected counts fell below 5 in any cell of the 2 × 2 table.

Power to detect replicated haplotypes as a function of OR and controls frequency was evaluated using the R method power.fisher.test running nsim = 10,000 simulations of the 2 × 2 haplotype-by-disease table, with the power defined as the fraction of tables with cases and controls haplotypes frequencies significantly different at the α = 1.0 × 10^−5^ (discovery) or 0.05 (replication) level (Supplementary Methods Section 8).

The prior steps were carried out using R version 3.6.1.

### Data availability statement

Accession of the UK Biobank (UKBB) data used in this study was approved under UKBB project #44891. Accession of the Discovery, Biology, and Risk of Inherited Variants in Breast Cancer (DRIVE) data used in this study was approved under dbGaP project #28544 and made through the NIH dbGaP (Study Accession phs001265.v1.p1).

TF-binding data in MCF-7 cells from ENCODE 3 (https://www.encodeproject.org/) (26,27) were downloaded through the UCSC Genome Browser (https://genome.ucsc.edu/index.html) (28) for the following factors: CTCF (experiment: ENCSR000DMR; bed file: ENCFF785NTC), ESRRA (experiment: ENCSR954WVZ; bed file: ENCFF541DRZ), FOXA1 (experiment: ENCSR126YEB; bed file: ENCFF160RLI), and MYC (experiment: ENCSR000DMQ; bed file: ENCFF370EQJ).

## Results

Exhaustive enumeration of combinations of SNPs is not an efficient way to look for genetic interactions. An alternative is to ask which patterns appear frequently in a dataset and test their association with a phenotype. In pursuit of the latter strategy, we performed two phases of Chromosome Overlap at three breast cancer GWAS hits to first identify haplotypes tagged by the GWAS hits (Phase 1) and then refine the haplotypes in a conditional analysis (Phase 2).

### Phase 1: Chromosome Overlap of regions containing a GWAS hit

For Phase 1, we selected regions that contained 50–60 genotyped SNPs within approximately 100 kb of (but not necessarily including) the GWAS hits: GRCh38 chr10:121,481,608– 121,680,765 (55 SNPs around rs2981578), chr11:69,419,318–69,616,860 (57 SNPs around rs554219), and chr16:52,486,414–52,645,181 (56 SNPs around rs4784227). See Supplementary Table S1 for lists of these SNPs. Starting with 18,022 UKBB chromosomes from 9,011 breast cancer cases, we formed all pairwise (*σ* = 2) overlaps in the initiation step and retained the following numbers of meta-chromosomes that showed evidence of breast cancer risk association and passed filtering by Fisher’s Exact Test with the indicated *P*-value thresholds: for the chromosome 10 region (hereafter abbreviated as “chr10”), 85 of 2,512,100 (*P* < 8.0 × 10^−21^); for chr11, 115 of 2,963,982 (*P* < 9.75 × 10^−10^); and for chr16, 245 of 3,459,313 (*P* < 1.0 × 10^−16^). These thresholds were chosen to prevent the total number of patterns at any iteration from reaching more than 300,000 (Supplementary Fig. S1; Supplementary Table S2). We iteratively formed meta-chromosomes by pairwise overlap, starting with the meta-chromosomes which passed filtering, and looked for unique patterns to be included in the next iteration; no additional filtering was applied. For chr11, the maximum number of unique patterns (285,829) was achieved at iteration 4, after which point there was a steady decline (Supplementary Fig. S1B); for chr10 and chr16, the maximum occurred at iteration 4 and 3, respectively (Supplementary Fig. S1A,C). At each step we determined the closed patterns by comparing the sets of meta-chromosomes before and after overlap and finding those that disappeared, i.e., did not appear in the newly formed set. After reaching the maximum, the number of closed patterns equaled the difference in the number of total patterns between subsequent iterations. Importantly, no new patterns appeared after the maximum, indicating that the algorithm could have terminated after three or four iterations. For example, the total number (286,393) of closed patterns found after 32 iterations for chr11 was equal to the number (27 + 77 + 160 + 300 + 285,829) of closed patterns that disappeared in iterations 1–4 plus the number remaining after iteration 4. See Supplementary Table S2 for the complete results.

Among the closed patterns found in each region, we expected to find haplotypes that were strongly linked to the GWAS hits and highly associated with breast cancer risk. The patterns that had the minimal likelihood ratio test (LRT) *P*-value for association with breast cancer risk were referred to as h1 (Table 1). Indeed, within our subset of UKBB women, each h1 was in strong linkage disequilibrium (LD) with its GWAS hit (h1-rs2981578: *r*^2^ = 0.70, *D*′ = 0.99; h1-rs554219: *r*^2^ = 0.78, *D*′ = 1.0; h1-rs4784227: *r*^2^ = 0.87, *D*′ = 1.0) and had a similar effect size (rs2981578[C]: HR = 1.25, *P* = 1.7 × 10^−51^; rs554219[G]: HR = 1.24, *P* = 3.0 × 10^−22^; rs4784227[T]: HR = 1.25, *P* = 1.2 × 10^−40^) and frequency. Note that rs4784227 on chr16 was the only GWAS hit genotyped in UKBB and part of h1; the other two GWAS-hit SNPs were not genotyped in UKBB but were imputed with high quality so that their effect sizes and LD properties could be measured.

**Table 1:** Frequency and breast cancer risk association of h1-subtypes in UKBB

| Variable | HR | LRT $P$-value<sup>a</sup> | Relative frequency / $10^{-4}$<br>(cases/controls)<sup>b</sup> |
| --- | --- | --- | --- |
| Chr10 |  |  |  |
| h1 | 1.30 | $1.0 \times 10^{-65}$ | 4,402/3,766 |
| rs2981578[C] | 1.25 | $1.7 \times 10^{-51}$ | 5,178/4,608 |
| LD<sup>c</sup>: $r^2 = 0.70$, $D' = 0.99$ | | | |
| h2 | 18.8 | $8.9 \times 10^{-11}$ | 6.659/0.2616 |
| h3 | 10.2 | $6.6 \times 10^{-9}$ | 7.768/0.5813 |
| h4 | 3.85 | $1.0 \times 10^{-7}$ | 18.31/4.825 |
| h5 | 10.5 | $2.2 \times 10^{-7}$ | 6.104/0.6852 |
| h6 | 6.29 | $2.5 \times 10^{-6}$ | 7.768/1.105 |
| h7 | 8.77 | $3.8 \times 10^{-6}$ | 5.549/0.4651 |
| Chr11 |  |  |  |
| h1 | 1.26 | $1.5 \times 10^{-21}$ | 1,165/947.1 |
| rs554219[G] | 1.24 | $3.0 \times 10^{-22}$ | 1,425/1,182 |
| LD<sup>c</sup>: $r^2 = 0.78$, $D' = 1.0$ | | | |
| h2 | 4.22 | $2.0 \times 10^{-7}$ | 14.44/3.372 |
| h3 | 5.87 | $3.2 \times 10^{-7}$ | 9.433/1.424 |
| h4 | 16.8 | $6.9 \times 10^{-6}$ | 3.329/0.1453 |
| Chr16 |  |  |  |
| h1 | 1.27 | $1.2 \times 10^{-41}$ | 2,550/2,118 |
| rs4784227[T] | 1.25 | $1.2 \times 10^{-40}$ | 2,808/2,367 |
| LD<sup>c</sup>: $r^2 = 0.87$, $D' = 1.0$ | | | |
| h2 | 50.4 | $1.7 \times 10^{-7}$ | 2.774/0.0 |
| h3 | 5.36 | $2.6 \times 10^{-7}$ | 10.54/1.773 |
| h4 | 7.49 | $1.9 \times 10^{-7}$ | 7.768/0.8720 |
| h5 | 3.22 | $1.6 \times 10^{-7}$ | 22.20/7.092 |
| h6 | 20.2 | $3.4 \times 10^{-7}$ | 3.884/0.1163 |
| h7 | 6.11 | $2.2 \times 10^{-6}$ | 7.768/1.105 |
| h8 | 2.68 | $2.7 \times 10^{-6}$ | 26.08/9.621 |
| h9 | 14.4 | $3.3 \times 10^{-6}$ | 3.884/0.1453 |
| h10 | 27.2 | $3.7 \times 10^{-6}$ | 2.774/0.02907 |
| h11 | 5.36 | $4.6 \times 10^{-6}$ | 8.323/1.453 |
| h12 | 6.41 | $7.0 \times 10^{-6}$ | 6.659/0.8720 |

<sup>a</sup> $P$ -value for the 1-d.f. likelihood ratio test between the model with h1 alone (resp. the null model) and the model with h1 and hx (resp. just h1 or the GWAS hit), adjusted for age, the first ten genotype principal components, and the remaining variables
<sup>b</sup> Among 18,022 case chromosomes and 344,046 control chromosomes
<sup>c</sup> Computed in UKBB cases and controls

The h1’s on chr10, chr11, and chr16 comprised 11, 19, and 26 non-contiguous SNP-alleles, most of which were the GRCh38 reference allele. Because closed patterns are the longest pattern shared by a group of chromosomes, it is possible that they contain a shorter pattern with the same support. Thus, we assessed whether some of the h1 alleles were unnecessary in specifying h1 and whether a reduced set of alleles could suffice. To test this possibility, we used rpart (24), the recursive partitioning method implemented in R, to derive three new sets of 10, 17, and 20 SNP-alleles, respectively, which had the same cases and controls frequencies as their longer versions. Because the shorter patterns were also subset patterns, and every chromosome with a superset pattern must have the subset, the reduced haplotypes were guaranteed to be perfectly correlated with their full-length versions. Henceforward, h1 refers to the reduced haplotype. The SNP-alleles of the three h1’s are given in Supplementary Table S5.

### Phase 2: Conditional analysis of h1-bearing chromosomes

Our original hypothesis was that rare haplotypes of large effect underlie the associations of GWAS hits with breast cancer risk. Specifically, the modest effect sizes of h1 and the GWAS hit we observed were due to a small subset of h1-bearing chromosomes associated with increased breast cancer risk along with many “false positive” haplotypes that carried the GWAS hit. To test this hypothesis, we applied Chromosome Overlap on h1-bearing case chromosomes in the topologically associating domains (TADs) containing the h1’s (see Figs. 2–4 below) whose boundaries were predicted using the 3D Genome Browser (25). We selected regions of the TAD, excluding the Phase 1 SNPs, which showed the greatest intensity of normalized Hi-C contacts on the hypothesis that risk haplotypes would physically interact with regulatory elements of nearby genes. For chr10 and chr11, the Phase 2 regions were directly upstream of h1, within approximately 55 and 10 kb of the Phase 1 regions, respectively: GRCh38 chr10:121,065,946–121,434,484 (103 SNPs); chr11:69,083,946–69,414,699 (92 SNPs). For chr16, the Phase 2 region spanned two sides of h1 and *CASC16*, removed by 150–200 kb from the Phase 1 region boundaries to capture a strong peak in the Hi-C matrix between the ends of the TAD (Fig. 4): chr16:52,168,409-52,313,907 and chr16:52,794,506–53,053,996 (30 + 72 = 102 SNPs). See Supplementary Table S3 for a complete list of the included SNPs. We were able to include more SNPs in Phase 2 than in Phase 1 because only a subset of case chromosomes carried h1: 7,934 chromosome 10’s, 2,100 chromosome 11’s, and 4,595 chromosome 16’s.

**Figure 2:**
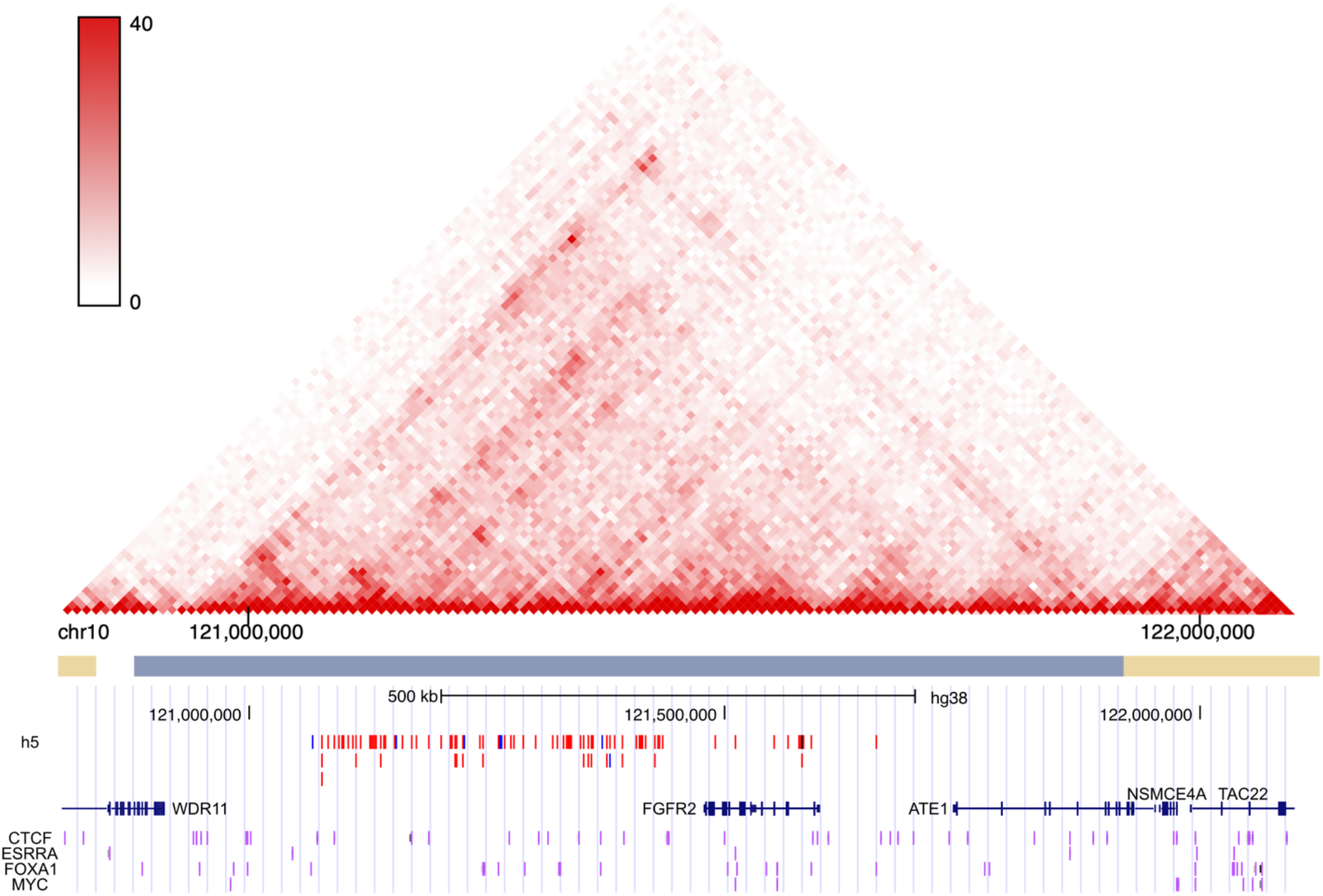
Haplotypes SNPs for replicated haplotypes at chromosome locus 10q26 (chr10). Top: Normalized HMEC Hi-C contact frequency and TAD boundaries from the 3D Genome Browser (25). Bottom: MCF-7 ChIP-seq peaks from ENCODE 3 (26,27) visualized in the UCSC Genome Browser (28). SNP tracks are shown in red and blue for reference and alternate alleles, respectively, and the GWAS hit rs2981578 (not part of the haplotype) is indicated in black. ChIP-seq for CTCF, ESRRA, FOXA1, and MYC are shown in purple. Genomic coordinates are GRCh38.

**Figure 3:**
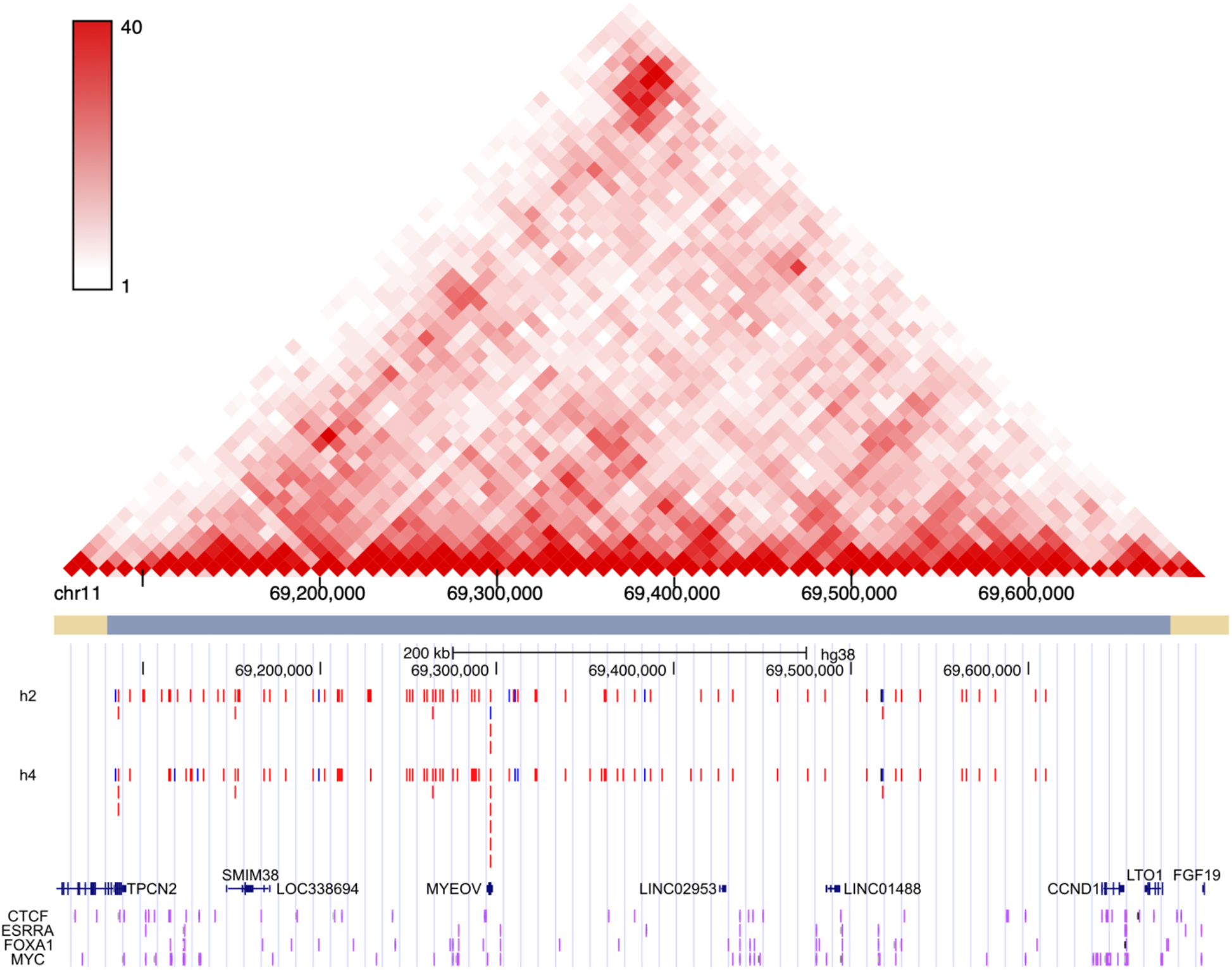
Haplotypes SNPs for replicated haplotypes at chromosome locus 11q13 (chr11). Normalized HMEC Hi-C contact frequency and TAD boundaries from the 3D Genome Browser (25). Bottom: MCF-7 ChIP-seq peaks from ENCODE 3 (26,27) visualized in the UCSC Genome Browser (28). SNP tracks are shown in red and blue for reference and alternate alleles, respectively, and the GWAS hit rs554219 (not part of the haplotypes) is indicated in black. ChIP-seq for CTCF, ESRRA, FOXA1, and MYC are shown in purple. Genomic coordinates are GRCh38.

**Figure 4:**
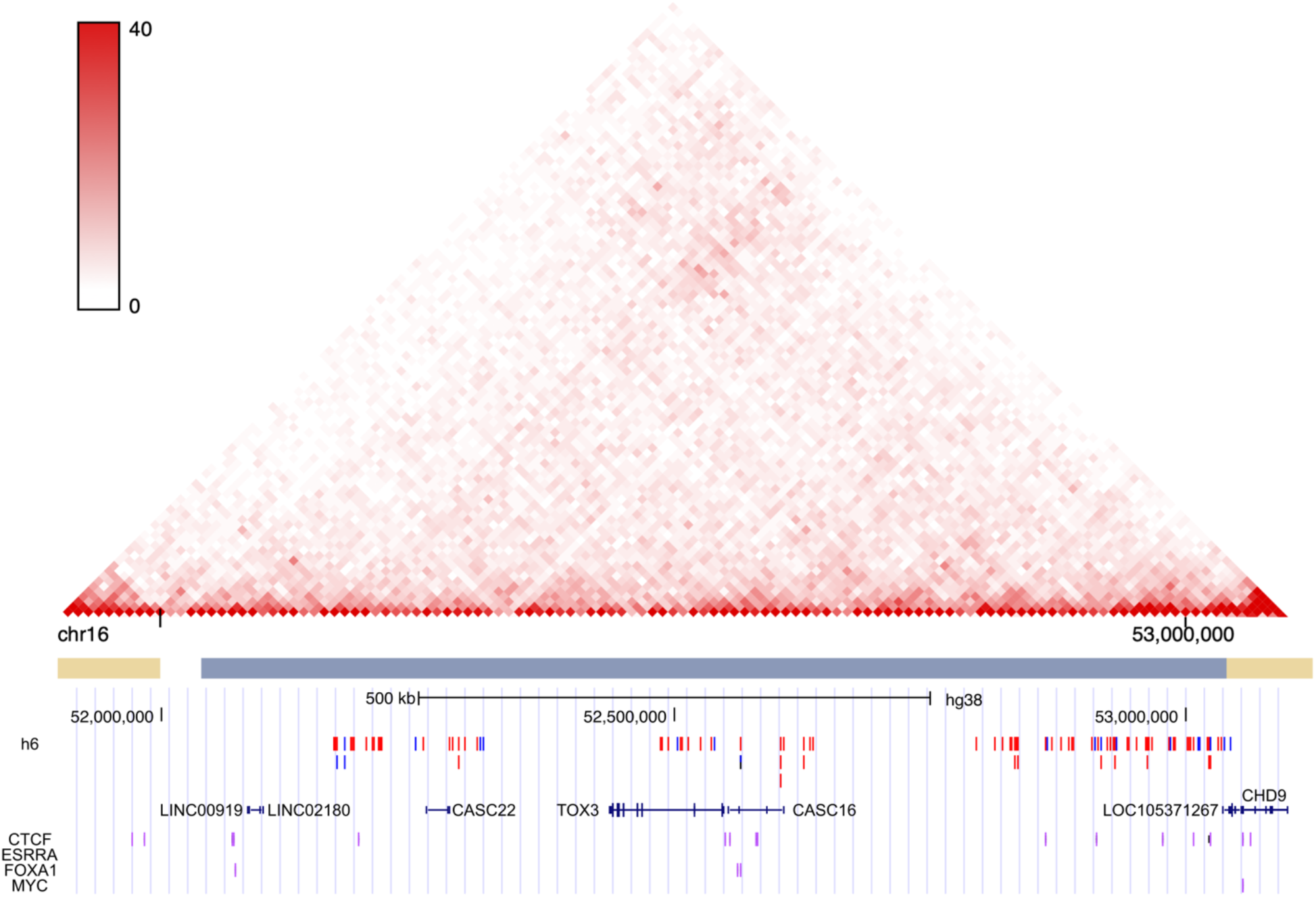
Haplotypes SNPs for replicated haplotypes at chromosome locus 16q12 (chr16). Normalized HMEC Hi-C contact frequency and TAD boundaries from the 3D Genome Browser (25). Bottom: MCF-7 ChIP-seq peaks from ENCODE 3 (26,27) visualized in the UCSC Genome Browser (28) SNP tracks are shown in red and blue for reference and alternate alleles, respectively, and the GWAS hit rs4784227 (part of the haplotype) is indicated in black/blue. ChIP-seq for CTCF, ESRRA, FOXA1, and MYC are shown in purple. Genomic coordinates are GRCh38.

After forming all pairwise overlaps during the initiation step, we found 7,887,179 unique patterns on chr10, 242,042 on chr11, and 5,136,876 on chr16. The difference in the number of patterns between chr11 and the other two chromosomes is owing to the difference of ten or eleven SNPs, illustrating the combinatorial complexity of the overlap process. Different association *P*-value thresholds (chosen to prevent the number of patterns from exceeding 400,000 at any iteration) were used to filter meta-chromosomes after the initiation step: 64 meta-chromosomes had *P* < 2.25 × 10^−6^ on chr10, 44 had *P* < 1.0 × 10^−4^ on chr11, and 33 had *P* < 6.0 × 10^−6^ on chr16. These were further filtered by removing patterns with identical cases/control frequencies and ORs to 39, 28, and 25, respectively, because Phase 2 was very sensitive to the number of starting patterns. We found a total of 397,819 closed patterns in 33 iterations on chr10; 167,998 in 25 iterations on chr11; and 368,806 in 24 iterations on chr16 (Supplementary Table S4).

Following overlap, we applied stepwise forward-selection to select an independent set of patterns associated with breast cancer risk using Cox regression. To assess the effect and independence of any new h1-subtype hx in the multi-haplotype model, we compared the model with h1 to the model with “h1 without hx” and “h1 with hx”; h1 was continually modified as new, statistically independent hx’s were added. We found several risk-associated patterns by this procedure: h2– h7 on chr10, h2–h4 on chr11, and h2–h12 on chr16 (Table 1; Supplementary Table S5). Each pattern was rare (controls frequency approximately 0.01–0.1%) and strongly risk-elevating (HR estimates in the range 2.68–50.4). We again tried to use recursive partitioning on h1-containing chromosomes to reduce the haplotypes but determined that the controls frequencies in the replication analysis (below) were better preserved by using full-length haplotypes (data not shown), which (except for h1) are used throughout.

To summarize the UKBB discovery analysis, we found six rare haplotypes on chr10, three on chr11, and eleven on chr16, which were all strongly associated with breast cancer risk.

### Replication in DRIVE

Next, we assessed whether the UKBB discovery-analysis findings could be replicated in an independent, large case-control study of breast cancer. For this analysis, we used 30,064 breast cancer cases and 25,282 controls in the DRIVE cohort. DRIVE haplotypes were imputed and phased at UKBB-genotyped SNPs using the TOPMed Imputation Server.

We first confirmed each of the three h1 associations with breast cancer risk in DRIVE using logistic regression, adjusting for age and the first ten genotype principal components (Table 2). For all three h1’s, an effect size and *P*-value similar to the UKBB results were observed. We next tested the Phase 2 results (h2–h7 on chr10, h2–h4 on chr11, and h2–h12 on chr16) individually by comparing the model with h1 alone to the model with h1 and hx (x = 2,…,12), both adjusted for age and the first ten genotype principal components. Since hx-bearing chromosomes also contained h1, h1 in the latter model was “h1 without hx” (as in the discovery analysis), allowing us to estimate the effect due to hx.

**Table 2:** Frequency and breast cancer risk association of h1-subtypes in DRIVE

| Variable | DRIVE OR | DRIVE $P$ | DRIVE relative frequency / $10^{-4}$ (cases/controls)<sup>c</sup> | Combined<sup>d</sup> OR | Combined<sup>d</sup> $P$ | Combined<sup>d</sup> relative frequency / $10^{-4}$ (cases/controls)<sup>e</sup> |
| --- | --- | --- | --- | --- | --- | --- |
| Chr10 |  |  |  |  |  |  |
| h1 | 1.26 | $1.1 \times 10^{-75a}$ | 4,354/3,812 | | | |
| rs2981578[C] | 1.22 | $7.3 \times 10^{-60a}$ | 5,239/4,755 | | | |
| LD<sup>f</sup>: $r^2 = 0.67$, $D' = 0.98$ | | | | | | |
| h2 | 0.656 | 0.23<sup>a</sup> | 0.9979/1.582 |  |  |  |
| h3 | 2.78 | 0.63<sup>b</sup> | 0.4989/0.1978 | 1.00 | 1.0<sup>b</sup> | 0.4989/0.5322 |
| h4 | 2.00 | 0.066<sup>a</sup> | 7.817/4.549 | 1.84 | 0.059<sup>a</sup> | 7.817/4.790 |
| h5 | 2.69 | 0.18<sup>a</sup> | 1.663/0.7911 | 4.22 | 0.024<sup>b</sup> | 1.663/0.6842 |
| h6 | 1.44 | 0.75<sup>a</sup> | 2.328/1.780 | 1.85 | 0.32<sup>a</sup> | 2.328/1.191 |
| h7 | 0.923 | 1.0<sup>b</sup> | 0.1663/0.1978 | 0.563 | 0.50<sup>b</sup> | 0.1663/0.4308 |
| Chr11 |  |  |  |  |  |  |
| h1 | 1.21 | $9.2 \times 10^{-22a}$ | 1,135/967.9 | | | |
| rs554219[G] | 1.21 | $3.2 \times 10^{-27a}$ | 1,464/1,252 | | | |
| LD<sup>f</sup>: $r^2 = 0.74$, $D' = 1.0$ | | | | | | |
| h2 | 2.13 | 0.022<sup>a</sup> | 8.648/4.351 | 2.17 | $1.8 \times 10^{-3a}$ | 8.648/3.497 |
| h3 | 1.07 | 0.68<sup>a</sup> | 4.490/4.351 |  |  |  |
| h4 | 11.9 | $2.7 \times 10^{-3a}$ | 2.162/0.1978 | 6.89 | $4.7 \times 10^{-8b}$ | 2.162/0.1520 |
| Chr16 |  |  |  |  |  |  |
| h1 | 1.22 | $1.1 \times 10^{-42a}$ | 2,431/2,093 | | | |
| rs478227[T] | 1.23 | $1.9 \times 10^{-50a}$ | 2,822/2,434 | | | |
| LD<sup>f</sup>: $r^2 = 0.82$, $D' = 1.0$ | | | | | | |
| h2 | >100 | 1.0<sup>b</sup> | 0.1663/0.00 | >100 | 0.13<sup>b</sup> | 0.1663/0.00 |
| h3 | 1.94 | 0.31<sup>a</sup> | 2.328/1.384 | 1.58 | 0.40<sup>a</sup> | 2.328/1.723 |
| h4 | 0.640 | 0.26<sup>a</sup> | 0.8316/1.582 | 0.578 | 0.16<sup>a</sup> | 0.8316/0.9630 |
| h5 | 0.971 | 0.31<sup>a</sup> | 7.151/7.713 | 1.02 | 0.37<sup>a</sup> | 7.151/7.172 |
| h6 | 2.89 | 0.38<sup>b</sup> | 0.6652/0.1978 | 3.13 | 0.022<sup>b</sup> | 0.6652/0.1267 |
| h7 | 0.343 | 0.020<sup>a</sup> | 0.6652/2.175 | 0.503 | 0.089<sup>a</sup> | 0.6652/1.242 |
| h8 | 1.51 | 0.34<sup>a</sup> | 8.482/5.933 |  |  |  |
| h9 | 1.08 | 1.0<sup>b</sup> | 0.1663/0.1978 | 0.924 | 1.0<sup>b</sup> | 0.1663/0.1520 |
| h10 | 0.988 | 1.0<sup>b</sup> | 0.1663/0.1978 | 2.51 | 0.35<sup>b</sup> | 0.1663/0.05068 |
| h11 | 1.12 | 1.0<sup>b</sup> | 0.8316/0.7911 | 1.04 | 0.76<sup>a</sup> | 0.8316/1.368 |
| h12 | 0.801 | 0.38<sup>a</sup> | 1.497/1.780 | 1.66 | 0.41<sup>a</sup> | 1.497/0.9883 |

<sup>a</sup> $P$ -value for the 1-d.f. likelihood ratio test between the model with h1 alone (resp. the null model) and the model with h1 and hx (resp. just h1 or the GWAS hit), adjusted for age and the first ten genotype principal components
<sup>b</sup> Fisher's Exact Test $P$ -value (unadjusted)
<sup>c</sup> Among 62,128 case chromosomes and 50,564 control chromosomes
<sup>d</sup> DRIVE and UKBB control chromosomes were only combined if the controls frequency difference was not statistically significant (Fisher's Exact Test $p \geq 0.05$ ; Table S6)
<sup>e</sup> Among 62,128 case chromosomes and 394,610 control chromosomes
<sup>f</sup> Computed in DRIVE cases and controls

We found that h2 and h4 on chr11 replicated directly in DRIVE with *P* < 0.05. The effect sizes of the replicated haplotypes were moderated in DRIVE but still larger than those of typical GWAS hits. It was also observed during the model-fitting process that genotype principal components had large and highly significant effects on breast cancer risk (Supplementary Table S7). Thus, we also computed the replication *P*-value by Fisher’s Exact Test, which is not adjusted for any genotype principal components, for haplotypes with expected counts less than 5 in any cell of the 2 × 2 table. No haplotype which satisfied this condition was directly replicated in DRIVE by this method.

We next asked whether the remaining haplotypes failed to replicate due to insufficient power. The DRIVE study has more breast cancer cases than UKBB but significantly fewer controls. If a haplotype frequency were elevated in cases relative to controls, our power to detect its effect would be increased if we had more controls who incidentally came from a population with similar haplotype frequency. To this end, we tested by Fisher’s Exact Test whether the DRIVE and UKBB controls frequencies were sufficiently similar to warrant our making a single group of combined controls: we pooled the controls when the differences were not statistically significant, i.e., *P* ≥ 0.05 (see Supplementary Table S6). The results of the model fitting are shown in Table 2 for the many haplotypes that met this criterion. We found that chr10 h5 and chr16 h6 (both with at least one expected cell count less than 5) replicated at *P* < 0.05 level by Fisher’s Exact Test. Taken together, one of six, two of three, and one of eleven rare haplotypes from the UKBB discovery analysis on chr10, chr11, and chr16, respectively, replicated at *P* < 0.05.

### Permutation analysis

We next asked whether replication of any of the rare haplotypes could have been expected by chance. To test this possibility, we repeated the Phase 2 analysis of chr11 using a permuted discovery dataset. Specifically, we permuted case/control status only among the 32,969 h1-carriers in UKBB such that the numbers of h1 homo- and heterozygotes were unchanged in each of the case and control groups. This step ensured that h1 had the same frequency and breast cancer association as the original unpermuted dataset; but any subtypes of h1 found in the (permuted) Phase 2 discovery analysis should have been associated with breast cancer only by chance and should not have replicated in DRIVE. We performed the Phase 2 analysis in the same manner as the original discovery analysis, including the same number (28) of filtered haplotypes kept in the original Phase 2 analysis for the remaining iterations. Then we assessed the replication of the discovered subtypes of h1 in the DRIVE data in the same manner as the original replication analysis, again combining controls when the DRIVE and UKBB controls frequencies were sufficiently similar (*P* ≥ 0.05). The results for a total of three permutations are shown in Table 3.

**Table 3:** Frequency and breast risk association of chr11 h1-subtypes in UKBB (permuted) and DRIVE

| Variable | HR/OR<sup>a</sup> | LRT P-value<sup>b</sup> | Relative frequency / 10<sup>-4</sup> (cases/controls)<sup>c</sup> | Combined<sup>d</sup> OR | Combined<sup>d</sup> P | Combined<sup>d</sup> relative frequency / 10<sup>-4</sup> (cases/controls)<sup>c</sup> |
| --- | --- | --- | --- | --- | --- | --- |
| Permutation 1 (50,058 closed patterns) |  |  |  |  |  |  |
| UKBB discovery |  |  |  |  |  |  |
| h2' | 4.58 | $1.4 \times 10^{-8}$ | 15.54/3.168 | | | |
| h3' | 6.18 | $3.8 \times 10^{-7}$ | 8.878/1.192 | | | |
| DRIVE replication |  |  |  |  |  |  |
| h2' | 1.35 | 0.68 | 5.821/4.549 | 1.21 | 0.92 | 5.821/3.345 |
| h3' | 1.23 | 0.93 | 9.480/7.911 |  |  |  |
| Permutation 2 (36,021 closed patterns) |  |  |  |  |  |  |
| UKBB discovery |  |  |  |  |  |  |
| h2'' | 5.45 | $4.6 \times 10^{-6}$ | 8.323/1.221 | | | |
| DRIVE replication |  |  |  |  |  |  |
| h2'' | 1.08 | 0.74 | 3.493/3.362 |  |  |  |
| Permutation 3 (23,169 closed patterns) |  |  |  |  |  |  |
| UKBB discovery |  |  |  |  |  |  |
| h2''' | 2.42 | $2.5 \times 10^{-6}$ | 34.40/13.52 | | | |
| DRIVE replication |  |  |  |  |  |  |
| h2''' | 1.12 | 0.61 | 16.80/15.43 | 1.12 | 0.67 | 16.80/13.76 |

<sup>a</sup> Hazard ratio for UKBB, odds ratio for DRIVE
<sup>b</sup> P-value for the 1-d.f. likelihood ratio test between the model with h1 alone and the model with h1 and hx, adjusted for age, the first ten genotype principal components, and (in UKBB only) the remaining variables
<sup>c</sup> Among 18,022 case and 344,046 control chromosomes in UKBB and 62,128 case and 50,564 control chromosomes in DRIVE
<sup>d</sup> DRIVE and UKBB control chromosomes were only combined if the controls frequency difference was not statistically significant (Fisher's Exact Test $P \geq 0.05$ )
<sup>e</sup> Among 62,128 case and 394,610 combined control chromosomes

Two notable observations can be made of the results of the permutation experiment. First, the UKBB discovery analysis could generate a small number (one or two) of haplotypes by chance alone. Second, none of the h1-subtypes discovered in the permuted UKBB data replicated by the LRT in DRIVE, suggesting the replication of h2 and h4 in the original analysis was likely valid.

### Features of the haplotypes

We next investigated the chromosomal locations and bioinformatics features of the SNPs of the replicated haplotypes. Figs 2–4 show the SNPs of the haplotypes in each chromosome region. Also included are HMEC Hi-C data from the 3D Genome Browser (http://3dgenome.org) (25) and ENCODE 3 (26,27) MCF-7 ChIP-seq data from the UCSC Genome Browser (http://genome.ucsc.edu) (28). The SNPs are not contiguous but lie in clusters throughout the TADs. The Hi-C data show that the 5′ region of the chr11 TAD interacts strongly with the 3′ region where *CCND1* is located (Fig. 3); haplotype SNPs are notably involved in the interaction. The chr10 TAD involves several interactions, although somewhat weaker than the strong peak on chr11. The chr16 TAD has the weakest interactions, although a broad peak between the ends of the TAD gave rise to several split haplotypes, only one of which replicated. As described above, because we pre-selected this region of the chr16 TAD for Phase 2, the absence of SNPs in other regions is not evidence of their not being involved in risk rare haplotypes. At the chr11 locus, we also observed at least four clusters of CTCF, ESRRA, and FOXA1 co-binding events (Fig. 3), suggesting that these transcription factors (TFs) may mediate the physical interactions. Importantly, this triplet of TFs is known to mediate cells’ response to estrogen (29,30). We also queried the HACER database (of Human ACtive Enhancers to interpret Regulatory variants) (31) to find other TFs binding enhancers of *CCND1* in MCF7 cells. A notable feature of this data was that all enhancers bound MYC, which also appeared in conjunction with the other three TFs (Fig. 3). The ChIP-seq data for these factors were sparser on chr10 and chr16 (Figs. 2 and 4).

## Discussion

Through the method developed in this paper, we have detected several rare haplotypes of large effect underlying GWAS hits associated with breast cancer risk. We discuss below (1) how our findings support a model of breast cancer risk driven by rare variants and genetic heterogeneity (13), (2) limitations of this study and alternative interpretations, and (3) methodological points regarding Chromosome Overlap.

Our findings in the vicinity of three strong GWAS hits support a model wherein rare haplotypes or mutations underlie the germline genetic risk for breast cancer associated with GWAS hits. Despite low power and a genetically heterogeneous replication population (see below), we discovered and replicated four of twenty rare haplotypes in the three chromosomal regions we examined. While each of the effect estimates was attenuated in the replication analysis, the DRIVE ORs of the replicated haplotypes were all 2.0 or larger, consistent with the twin data supporting a single high-risk variant’s being the cause of familial aggregation of breast cancer (6); in contrast, each GWAS hit was associated with increased risk by a factor of approximately 1.2. Although these rare haplotypes alone do not fully capture the association of each of the GWAS hits with breast cancer risk, when pooled with other rare haplotypes they might. We estimate that chr11 haplotypes h2 and h4 contribute an amount 2*p*(1 − *p*)*β*^2^ = 0.00050 and 0.00024, respectively, to the narrow-sense frailty-scale heritability, where *p* is the frequency in replication controls and β is the logarithm of the effect size (1,2), in comparison with 0.0080 for the much more common GWAS hit rs554219. But power calculations (Supplementary Methods Section 7; Supplementary Table S9) suggest that at least 16 and 5 haplotypes with similar respective frequencies and effect sizes to h2 and h4 should exist, which together account for more than 100% of the tagged heritability. If genuine, these haplotypes represent either (a) approximations of single risk-elevating mutations or (b) true (or approximate) risk-elevating haplotypes with interacting combinations of alleles. Our study was not able to distinguish these two possibilities, but various plausible scenarios are conceivable that explain the results in terms of hypotheses (a) or (b).

According to quantitative evolutionary theory (9), mutations arise on ancestral haplotypes in proportion to their frequency in the population, so that the rarer the haplotype, the stronger its correlation with the mutation. If the risk mutation arose only recently, the rare haplotype on which it arose would be unbroken by recombination and associated with disease risk. In contrast, if the mutation is old, only a short region of the haplotype about the mutation will be shared among affected individuals. A short segment is necessarily more common than a long one, so the risk attributed to it (or to common variants it comprises) will eventually be diluted by many “false-positive” non-mutation carriers. Paradoxically, as the region of the mutation becomes more precisely pinpointed, the risk attributed to it decreases. Thus, it appears that Chromosome Overlap is designed to specifically detect recent mutations that arose on rare haplotypes. However, our detecting a rare haplotype in each of three regions seems *a priori* unlikely, because it is on common haplotypes that mutations tend to arise. On this argument, the Chromosome Overlap strategy appears destined to fail. However, an important caveat to this deduction is that any haplotype is rare if it is long enough. Indeed, each replicated haplotype we found was long (spanning 0.5–1 Mb of a TAD) and constituted of independent, individually common blocks of SNPs in strong LD (determined using the R package Big-LD (32) in Supplementary Methods Section 7; Supplementary Table S8; Supplementary Figs. S3–S5). These haplotypes cannot be too old, or else the only shared segment will be a short (and therefore likely common) region containing the mutation. These considerations imply that the haplotypes detected by Chromosome Overlap carry relatively recent mutation events, with the stronger results on chr11 likely indicating more recent risk mutations resulting in extensive haplotype sharing and the weaker results on chr16 being due to older mutations with less haplotype sharing. A rare-haplotype model such as this would help explain why GWAS hits are poor approximations for rare causal variation.

It is, however, also possible that the rare haplotypes do not represent single risk mutations, but rare combinations of alleles whose interaction predisposes to disease. Few rare single variants have been found to explain synthetic associations of GWAS hits at breast cancer loci (11,12). Furthermore, despite the prevalence of GWAS hits, comparatively few rare mutations have been found in family-based linkage studies of breast cancer with subsequent positional cloning (33,34). The alternative that explains these observations is a preponderance of *cis* interactions of variants on the same chromosome. These allele combinations would not be detected easily by linkage analysis if they are rare or not frequently shared across case families; they are therefore not inconsistent with the general absence of linkage-analysis discoveries in regions with GWAS signals that are predicted to exist under the synthetic-associations model (8,10). Neither is the causal-haplotype hypothesis inconsistent with the finding that genetic epistasis—the interaction between genes—makes a relatively minor contribution to heritable risk (35,36), because previous studies that have looked at variant combinations (17-19,35) have considered *trans* interactions between chromosomes. The ChIP-seq and Hi-C data, especially on chr11, suggest that the whole of the haplotype may be involved in altering disease risk. For example, the high-intensity of Hi-C contacts on chr11 are complemented by ChIP-seq peaks for CTCF, FOXA1, and ESRRA (Estrogen Related Receptor Alpha). The estrogen receptor (ER) binds throughout the genome at enhancers distant from the start sites of genes it regulates (37,38), including *CCND1*. Several proteins are involved its recruitment, including the pioneer factor FOXA1 (29,38) and CTCF which acts upstream of FOXA1 to drive ER-mediated transcription via chromosome loops (39) and partitions the genome into ER-responsive blocks (40). A small fraction of binding events genome-wide involve all three factors which then contribute to down-regulation of estrogen target-genes (29,30). A fourth TF with frequent binding in the *CCND1* TAD is coded by the proto-oncogene *MYC*. MYC-binding was identified by computational analysis in a *CCND1* enhancer encompassing the original GWAS hit rs614367 (31), later replaced with rs554219 using fine-mapping (22), and it has long been known that MYC can repress *CCND1* expression (41,42). As certain SNPs in the haplotype involve simultaneous binding of all these factors, suppression of CCND1 may be the high-risk event leading to breast cancer that is captured by the rare haplotypes. On this hypothesis, a rare, unfortunate series of recombination events that juxtaposes key alleles results in a risk haplotype and the disease; different families could segregate the same risk haplotype through recombination despite its not being inherited from a common ancestor. Although we observed the strongest evidence of consequential TF-binding around rs554219 on chr11, others have demonstrated robust, allele-specific interactions between rs4784227 and a SNP in the *TOX3* promoter on chr16, possibly involving FOXA1 (43), and OCT4 and RUNX2 binding have been implicated in mediating the effect of rs2981578 on chr10 (21).

The generalizability of our results to other GWAS hits needs to be investigated. An alternative interpretation of our findings is that the haplotypes we identified reflect small but direct effects of the GWAS hits (as per the polygenic model) and that the large hazard ratios of the rare haplotypes we found are statistical artifacts. Using reporter assays, other studies have found that alternate alleles of the GWAS hit at each of the three loci can cause changes in gene expression or TF-binding (21-23,43). Against this interpretation, a non-*BRCA1*/*2* family from the Netherlands with six cases of breast cancer was found to have a strong linkage peak at the 11q13 locus as well as in a chromosome 14 region containing the *FOXA1* gene (44). The strength of this effect is inconsistent with the small OR of the GWAS hit, and the involvement of another gene known to be important in CCND1 regulation (i.e., *FOXA1*) suggests that a specific pathway may be driving disease risk. There are two points that need to be considered when weighing the evidence for these two interpretations. First, the women in our DRIVE analysis, although all of European ancestry, are genetically more diverse than their counterparts in our UKBB analysis. Genetic diversity has limited the generalizability of results from previous linkage studies of breast cancer (33,34), and in fact, we observed strong and significant correlations between genetic ancestry PCs and breast cancer risk in DRIVE (even after selecting the European-ancestry subset (Fig. S2B)) that were not present in the more homogeneous UKBB “white British” sample (Supplementary Table S7). Second, an important limitation of our study was low power. For example, the estimated power to discover in UKBB a haplotype like chr11 h2 was only 6% (Supplementary Methods Section 7; Supplementary Table S9), suggesting that 16– 17 such haplotypes may exist that could not be detected. Similarly, we had only 17% power to discover (in UKBB) rarer haplotypes like chr11 h4 with higher replication ORs, suggesting that 5–6 similar ones may exist (Supplementary Table S9). Finally, because our study was limited to UKBB-genotyped SNPs, there may be many other risk haplotypes in the population that we could not observe or measure. In summary, while the reporter-assay evidence is supportive of the polygenic model, it does not refute the genetic heterogeneity and multiple-rare-variant/haplotype basis of breast cancer risk. On the other hand, the evidence from linkage analysis as well as large twin/family studies supports the latter model and is inconsistent with the polygenic interpretation.

Chromosome Overlap is a complementary method to bottom-up closed-pattern miners like LCM (16) and has the potential for further improvement. LCM and Chromosome Overlap share similar limitations when the number of SNPs becomes large. Relator *et al*. had to split thousands of genes into several blocks of 250 in order to find closed gene-expression patterns by LCM among hundreds of breast- and ovarian cancer patients (19). Chromosome Overlap is also limited in the number of SNPs it can handle, but it differs from LCM in that the longest patterns are found first; LCM appears to take longer to find these patterns, especially in large datasets, because it only finds them after mining shorter patterns. While we used pattern disappearance as the criterion to ensure that we captured all closed patterns, we found empirically that most, if not all, closed patterns appeared within the first few iterations. If this observation can be theoretically justified, or if we are willing to forfeit the patterns of later iterations, denser genotype data and longer regions can be studied in the future by Chromosome Overlap. Currently, the initiation and filtering steps of Chromosome Overlap knowingly reject some patterns in order to capture the most likely candidates in reasonable time, but by increasing the value of *σ* it may be possible to capture more of them. Finally, Chromosome Overlap is fully parallel, and with enough computing nodes, the overlaps can be computed in a relatively short time.

In conclusion, through a new closed-pattern-mining method called Chromosome Overlap, we discovered and replicated several rare, risk haplotypes underlying three breast cancer GWAS hits, with the possibility that there may be many more. Similar to the synthetic-associations and genetic-heterogeneity hypotheses, these results suggest that GWAS hits derive their signal by averaging over many “false positive” haplotypes and artificially smooth the risk gradient across women. If replicated in the vicinity of other GWAS hits and backed up by functional studies, the rare-haplotype hypothesis may challenge the prevailing polygenic, additive model of breast cancer risk.

## Data Availability

All data produced in the present study are available upon reasonable request to the authors

https://www.ukbiobank.ac.uk/

https://www.ncbi.nlm.nih.gov/projects/gap/cgi-bin/study.cgi?study_id=phs001265.v1.p1

## Acknowledgments

The authors thank the high-performance computing facility at St. Jude Children’s Research Hospital for computational support.

