## Supplementary Information for "Potential Misrepresentation of Inherited Breast Cancer Risk by Common Germline Alleles"

### List of Figures

Supplementary Figure S1 The total (left) and closed (right) patterns found at each iteration of Phase 1 at 10q26 (chr10) (A), 11q13 (chr11) (B), and 16q12 (chr16) (C). 15

### 1. Chromosome Overlap

In the next three subsections we introduce the concepts and notation required to describe the three steps of Chromosome Overlap.

##### *1.1 Initiation*

The $j^{\text{th}}$ chromosome in a sample is a column vector $\mathbf{x}_j = (x_{1,j}, \ldots, x_{m,j})^T$ of alleles $x_{k,j} = S_k$ or $s_k$ at $m$ biallelic SNPs, with uppercase and lowercase denoting the GRCh38 reference and alternate alleles, respectively. Overlap of a number $\sigma$ of chromosomes produces a meta-chromosome representing the set's maximal shared pattern (for this paper, $\sigma = 2$ because we consider only pairs of chromosomes). Meta-chromosomes in general have fewer alleles than do each of the chromosomes they comprise and can be themselves overlapped to produce even shorter meta-meta-chromosomes (Supplementary Methods section 1.3; Main Text Fig. 1C). The number of meta-chromosomes generated grows combinatorically with the number $\sigma$ of overlapped chromosomes and the total number of chromosomes under study (i.e., twice the number $N$ of study subjects). Because each overlap is straightforward to compute (Main Text Fig. 1A), the discovery of closed patterns is amenable to parallel implementation.

There are two keys to efficiently performing all $\sigma$-overlaps in parallel. The first is recognizing that, because all chromosomes are of equal length, they can be represented in an $m \times 2N$ (transposed) haplotype matrix arranged with $m$ SNPs down the rows and $2N$ chromosomes from $N$ cases across the columns (Fig. 1A). Then each computing node can evaluate the overlap of a fixed set of chromosomes with every other. Formally, let the $j^{\text{th}}$ chromosome be represented by the $j^{\text{th}}$ column $\mathbf{x}_j$ of the matrix. In general, $\sigma - 1$ chromosomes $\mathbf{x}_{i_1}, \ldots, \mathbf{x}_{i_{\sigma-1}}$ (where $i_1 < \cdots < i_{\sigma-1}$) are *fixed* and overlapped singly with every other *variable* chromosome $x_{i_\sigma}$ satisfying $i_\sigma = j > i_{\sigma-1}$. The row numbers $k$ at which $x_{k,i_1} = \cdots = x_{k,i_\sigma}$ and the allele value (0 or 1) are output as a list $(k_1, x_{k,i_1}), (k_2, x_{k,i_2}), \ldots, (k_\ell, x_{k,i_\ell})$ at every column $j > i_{\sigma-1}$ of the matrix. This list is a new pattern called a meta-chromosome. A single operation is complete when all $2N - i_{\sigma-1}$ meta-chromosomes comprising $\sigma$ chromosomes each are generated for a single set of $\sigma - 1$ fixed chromosome(s). Each node computes a fixed range of operations.

The second key to parallelization is that, while we know how many $\sigma$-combinations of chromosomes there are, we do not immediately know which chromosomes belong to a specific combination, e.g., the 1,000$^{\text{th}}$. We overcome this difficulty by using an algorithm that uniquely indexes each combination of $\sigma$ chromosomes in a triangular array (Supplementary Methods section 2; Main Text Fig. 1B). The chromosomes in the $\mathsf{L}^{\text{th}}$ tuple can be quickly accessed without exhaustive enumeration of all tuples, and each computing node can be instructed to evaluate a fixed number of overlaps starting from the $\mathsf{L}^{\text{th}}$. For the same haplotype matrix, a range of the $\binom{2N}{\sigma-1}$ operations is determined using the triangular array (Main Text Fig. 1B). Although for pairwise overlap ($\sigma = 2$), splitting up $\binom{2N}{\sigma-1} = 2N$ combinations of $2N$ fixed chromosomes is straightforward, we need the triangular array below (Supplementary Methods section 1.3) during the iteration steps when combinations of meta-chromosomes are formed.

Finally, the lists of meta-chromosomes from each node are combined and removed of duplicates. Initiation is represented by the first column of Main Text Fig. 1C.

##### *1.2 Filtering*

After the initiation step, there are up to $\binom{2N}{\sigma}$ distinct meta chromosomes. If all meta-chromosomes are repeatedly paired and overlapped (see Supplementary Methods section 1.3, below), there will quickly become an astronomical number of them. To limit the number of patterns considered, the potential of each to generate a risk-associated pattern downstream is evaluated using Fisher's Exact Test for breast cancer association; those which do not pass with a pre-specified statistical-significance ($P$-value) threshold are deemed low-potential and discarded. Although this step prevented the discovery of patterns whose all longest subsets did not pass the statistical-significance threshold, we reasoned that patterns with the strongest effects on breast cancer risk would show some level of statistical significance after the initiation step and that iteration would increase their significance. Typically, we adjusted the statistical-significance threshold so that the maximum number of meta-chromosomes (reached in only three or four iterations) generated in any iteration did not exceed 300,000 (Phase 1) or 400,000 (Phase 2).

An additional filtering step was applied when the primary filtering was not enough to prevent the overlap process from blowing up. Specifically, we selected the shortest pattern from each set of patterns that, while not related as subset-superset, had the same frequency and association OR/$P$-value from Fisher's Exact Test.

##### *1.3 Iteration*

Once the meta-chromosomes found in the initiation step have been filtered, the meta-chromosomes are themselves iteratively overlapped (second and third columns of Main Text Fig. 1C).

Because meta-chromosomes are not all the same length, they cannot be arranged in a matrix like chromosomes can. Furthermore, the restriction in the initiation step that variable chromosomes satisfy $i_\sigma > i_{\sigma-1}$ results in an unequal distribution of labor, as some nodes evaluate a smaller number $2N - i_{\sigma-1}$ of columns than others; this problem is only magnified when working with the much longer lists of meta-chromosomes. Instead, each unique combination of meta-chromosomes is directly indexed, and the triangular array (Main Text Fig. 1B) is used to assign a range of index combinations to each computing node without explicit enumeration. To perform the overlap operation, a node iterates through the SNP-alleles of the $\sigma^{\text{th}}$ meta-chromosome in the combination and chooses the ones that appear in each of the prior meta-chromosomes $i_1, \ldots, i_{\sigma-1}$; this approach is possible because the longest shared pattern is, by definition, a subset of any one of the meta-chromosomes. The output from each node is a list of meta-meta-chromosomes, which are then pooled and removed of duplicates before the next round; no additional filtering is done.

The importance of iteration with $\sigma = 2$ is that it discovers all closed patterns among the set of meta-chromosomes present after the initiation and filtering steps (i.e., the set of filtered meta-chromosomes). Because we start with filtered meta-chromosomes, we cannot even form all combinations of the chromosomes they comprise; the best we can do is find all closed patterns among the filtered set. A proof that Chromosome Overlap does exactly this is given in Supplementary Methods section 3.

Closed patterns are identified during the iterations in the following way. A meta-chromosome that appears for the first time as the output of some overlap operation must be closed. Alternatively, if a meta-chromosome is a proper subset of another, the result of overlap of the two is the meta-chromosome itself: it "survives" the iteration. However, if a meta-chromosome is a subset of no other meta-chromosome, it will disappear from the list of meta-meta-chromosomes in the next iteration. Once all patterns that have appeared have subsequently disappeared, no more non-null overlaps can be formed, and thus all closed patterns have been found. At the end of each iteration the lists of meta-chromosomes from before and after the overlap operations are compared to determine which meta-chromosomes have disappeared. Although it is possible for a pattern to appear at a late iteration, in practice, most are formed within four additional rounds.

#### 2. Triangular-array algorithm

In this section we describe the algorithm for determining the chromosome combination corresponding to an arbitrary index. As in the main text, $N$ is the number of samples and $\sigma$ is the degree of overlap. Our problem is to find, for a given $N$ and $\sigma$, what chromosome combination is the $1000^{\text{th}}$ (or any other number)? In order to answer this question without generating a list of all $\binom{2N}{\sigma}$ chromosome combinations, we first index each combination by $\mathsf{L} \in \left\{0, 1, \ldots, \binom{2N}{\sigma} - 1\right\}$ corresponding to a unique multiindex $\mathsf{I} = i_1, i_2, \ldots, i_\sigma$ of $\sigma$ different chromosomes with $i_1 < i_2 < \cdots < i_\sigma$. The mapping from $\mathsf{L}$ to $\mathsf{I}$ is accomplished iteratively using the following lemma. It what follows, we put $N \leftarrow 2N$ for notational convenience.

**Lemma 1.** *For any integers $N$ and $\sigma \leq N$,*

$$\binom{N}{\sigma} + \binom{N-1}{\sigma} + \cdots + \binom{N-(N-\sigma)}{\sigma} = \binom{N+1}{\sigma+1}.$$

*Proof.* We will prove the lemma by induction. Clearly the result holds when $\sigma = 1$ as the usual integer-summation formula, and the result when $\sigma = N$ is trivial. Now suppose that each term on the l.h.s. is the number of ways to arrange $N - j$ things $(0 \geq j \geq N - \sigma)$ in a $\sigma$-dimensional triangular array. Thus, $\binom{N}{\sigma}$ corresponds to the number of $\sigma$-tuples of the items $\{2, \ldots, N+1\}$, $\binom{\sigma}{\sigma}$ to the $\sigma$-tuples of $\{N - \sigma + 2, \ldots, N + 1\}$, and, in general, $\binom{N-j}{\sigma}$ to $\sigma$-tuples of $\{j + 2, \ldots N + 1\}$. Now, append to each tuple an element $i_1$ which is one less than the minimum item $j+2$ in the $\sigma^{\text{th}}$ dimension of each array. In this way we form all $\sigma + 1$-combinations of the elements $\{1, 2, \ldots, N + 1\}$ in a $\sigma + 1$-dimensional array, of which there are $\binom{N+1}{\sigma+1}$. Because the l.h.s. is equal to the r.h.s., the result follows for all $\sigma$ up to $N$. $\square$

To use Lemma 1, arrange the values of $\mathsf{I}$ in a $\sigma$-dimensional triangular array such that $i_1$ takes values in $\{1, \ldots, N - \sigma + 1\}$, $i_2$ takes values in $\{i_1 + 1, \ldots, N - \sigma + 2\}$, and in general $i_j$ takes values in $\{i_{j-1} + 1, \ldots, N - \sigma + j\}$ (see Figure 1B in the main text). Following the convention that the first dimension of a matrix is the rows, we have that $i_1$ corresponds to the $\sigma^{\text{th}}$ dimension, $i_\sigma$ the first, and $i_j$ the $\sigma - j + 1^{\text{th}}$. Starting from $i_1 = 1, i_2 = 2, \ldots, i_\sigma = \sigma$ and incrementing from right to left (i.e., ranging through all values in the first dimension before changing the value in the second), the $\mathsf{L}^{\text{th}}$ element will be the multiindex $\mathsf{I}$. Determining the indices $i_j$ is equivalent to determining the value or *layer* of the $j^{\text{th}}$ dimension of the array in which the $\mathsf{L} = \mathsf{L}^{(0)\text{th}}$ element resides. To determine the first layer, recognize that, for fixed $i_1$, there are only $\binom{N-i_1}{\sigma-1}$ chromosome combinations available to the remaining indices, because $\forall j > 1$, $i_j > i_1$. Then $i_1$ is determined by subtracting layers in the $\sigma^{\text{th}}$ dimension until the smallest $k$ is found which satisfies $\mathsf{L}^{(0)} - \sum_{\ell=1}^{k} \binom{N-\ell}{\sigma-1} < 0$, or equivalently (by Lemma 1) $\varepsilon_1 := \left[\binom{N}{\sigma} - \binom{N-k}{\sigma}\right] - \mathsf{L}^{(0)} > 0$, and putting $i_1 \leftarrow k$. The error $\varepsilon_1$ is the remainder beyond $\mathsf{L}^{(0)}$ in the $\sigma - 1$-dimensional array, which has $\binom{N-i_1}{\sigma-1}$ combinations. Thus, after determining $i_j$, put $\mathsf{L}^{(j)} \leftarrow \binom{N-i_j}{\sigma-j} - \varepsilon_j$ and get the position of the multiindex $i_{j+1}, \ldots, i_\sigma$ in the $\sigma - j$-dimensional array of $\binom{N-i_j}{\sigma-j}$ combinations. To do so, solve again for the smallest $k$ such that $\varepsilon_{j+1} := \left[\binom{N-i_j}{\sigma-j} - \binom{N-i_j-k}{\sigma-j}\right] - \mathsf{L}^{(j)} > 0$ and put $i_{j+1} \leftarrow i_j + k$, because $i_{j+1}$ starts from $i_j + 1$. The algorithm terminates when $\mathsf{L}^{(\sigma-1)} = \binom{N-i_{\sigma-1}}{1} - \varepsilon_{\sigma-1}$ is the position of the multiindex $i_\sigma$ in the 1-dimensional array of $N - i_{\sigma-1}$ "combinations" of single chromosomes.

#### 3. Completeness of Chromosome Overlap

Here we establish the result that Chromosome Overlap with $\sigma = 2$ finds all closed patterns. First we need the

**Lemma 2.** *$\sigma$-overlap of meta-chromosomes each comprising $\sigma + k$ ($k \geq -1$) chromosomes can be accomplished with as few as $\sigma + k + 1$ distinct chromosomes.*

*Proof.* We begin by constructing a table of $x$ vs. $y$ in which each entry $(x, y)$ is defined as the smallest integer $n$ satisfying

$$x \leq \binom{n}{y}, \tag{1}$$

as in

| $x$ \ $y$ | 1 | 2 | 3 | 4 | 5 | 6 |
|---|---|---|---|---|---|---|
| 1 | 1 | 2 | 3 | 4 | 5 | 6 |
| 2 | 2 | 3 | 4 | 5 | 6 | 7 |
| 3 | 3 | 3 | 4 | 5 | 6 | 7 |
| 4 | 4 | 4 | 4 | 5 | 6 | 7 |
| 5 | 5 | 4 | 5 | 5 | 6 | 7 |
| 6 | 6 | 4 | 5 | 6 | 6 | 7 |

Thus, $n$ is the minimum number of distinct chromosomes in the overlap of $x$ meta-chromosomes each comprising $y$ chromosomes. That is, if there are $n$ distinct chromosomes, $\binom{n}{y}$ groups of size $y$ can be formed; overlap of $x$ such groups can be accomplished using only $n$ chromosomes if Eq. (1) is satisfied, resulting in an $n$-tuple meta-meta-chromosome. The table can be built recursively by noting that $(1, y) \to y$ (i.e., a single meta-chromosome comprises exactly $y$ chromosomes) and then deriving $(x, y)$ from $(x-1, y) \to n$ as either $n$ if Eq. (1) holds or $n + 1$ if not.

It is easy to see that Eq. (1) is satisfied by $n = y + 1$ when $x \leq y + 1$, because $\binom{y+1}{y} = y + 1$. If also $x > 1$, then $n = y + 1$ is minimal, because only $x = 1$ satisfies $x \leq \binom{y}{y}$. Thus, rows $x = 2$ to $x = y + 1$ of column $y$ of the table will all be $(x, y) \to y + 1$. (It may also be noted that $(x, y)$ can be derived recursively from $(x, y - 1) \to n = y$ as $n + 1 = y + 1$, for rows $x = 2$ to $x = (y - 1) + 1 = y$.) The lemma follows by putting $\sigma \leftarrow x$ and $\sigma + j \leftarrow y$: because $\sigma + k \geq \sigma - 1$, the minimum number of distinct chromosomes needed for $\sigma$-overlap of $\sigma + k$-tuples is $(\sigma, \sigma + k) \to \sigma + k + 1$. □

We will now use Lemma 2 to prove that is always possible to add any single chromosome to each meta-chromosome at each iteration. Recall that we may start the iterations at the initiation step or after filtering. While meta-chromosomes technically are overlapped to become meta-meta-chromosomes, we will use the term "meta-chromosome" more generally to mean the result of overlap of $\sigma$ chromosomes or meta-chromosomes at any iteration $k \geq 0$. A meta-chromosome then comprises $\sigma$ (less-highly overlapped) meta-chromosomes or some greater number of (filtered meta-)chromosomes, and it is convenient to refer to any one as the list $j_1 j_2 \dots j_n$ of $n$ (filtered meta-)chromosomes that have been overlapped to form it. Now we may state the

**Theorem 3.** *At the end of iteration $k \geq 0$ (or the beginning of iteration $k+1$) of iterated overlap, all $\sigma + k = \sigma + (k-1) + 1$-tuples of $N$ (filtered meta-)chromosomes are formed.*

*Proof.* We will prove this result by induction. Clearly all $\sigma$-tuples are formed at the end of iteration $k = 0$, as each meta-chromosome now comprises $\sigma$ (filtered meta-)chromosomes. Then, together with Lemma 2, starting from all $\sigma$-tuples at the beginning iteration $k = 1$, it will be possible to form by $\sigma$-overlap any meta-chromosome comprising $\sigma + 1 = \sigma + (1-1) + 1$ (filtered meta-)chromosomes at the end of it, as every $\sigma$-tuple is available to supply one of the $N$ (filtered meta-)chromosomes that another $\sigma$-tuple does not contain. Thus, all $\sigma + 1$ tuples are formed at the end iteration $k = 1$ (or the beginning of iteration $k + 1 = 2$). Now assume that all $\sigma + k$-tuples have been formed at the end of iteration $k$ (or the beginning of iteration $k + 1$). Again by Lemma 2, $\sigma$-overlap of $\sigma + k$-tuples can be accomplished with as few as $\sigma + k + 1$ (filtered meta-)chromosomes. And since all $\sigma + k$ tuples are available, any and all $\sigma + k + 1 = \sigma + (k + 1 - 1) + 1$-tuples can be formed at the end of iteration $k + 1$, thus proving the induction hypothesis.

One case remains to be considered. Overlaps which produce a null meta-chromosome are not overlapped further. But observe that adding any (filtered meta-)chromosome to a null meta-chromosome still results in a null meta-chromosome. Thus, the null meta-chromosome represents all $\sigma + k$-tuples at the end of iteration $k$ which produced a null overlap, which are therefore available to produce null meta-chromosomes in iteration $k + 1$. In other words no $\sigma + k + 1$-tuple has been missed by our only overlapping non-null meta-chromosomes. □

Theorem 3 shows that iterated $\sigma$-overlap produces all $\sigma$-tuples at the end of iteration $k = 0$, all $\sigma + 1$ tuples at the end of iteration 1, all $\sigma + 2$ tuples at the end of iteration 2, etc. This pattern is contrary to the naive assumption that $\sigma^2$-tuples are formed from $\sigma$-tuples at the end of iteration 1 and $\sigma^{k+1}$-tuples from $\sigma^k$-tuples at the end of iteration $k$. The reason is that when $\sigma$ meta-chromosomes share some of their $\sigma$ (filtered meta-)chromosomes (as 12 and 13 do), their overlap (123) will include fewer than $\sigma^2$ distinct objects. Since it is always possible to add to any meta-chromosome a single (filtered meta-)chromosome that it does not contain, Chromosome Overlap with $\sigma = 2$ discovers all closed patterns. Although overlap with $\sigma > 2$ skips meta-chromosomes with fewer than $\sigma$ (filtered meta-)chromosomes (and is therefore not complete), it may still be a practical strategy to retain meta-chromosomes which otherwise would have been filtered out after the initiation step. $\sigma > 2$ was not investigated in this study but is still an option in the program code.

#### 4. Chromosome Overlap run specifics

Supplementary Table S1 gives the SNPs selected in each region for Phase 1. All coordinates are GRCh38.

**Supplementary Table S1**: SNPs included in Phase 1

| rsid | Position | Reference allele | Alternate allele |
|---|---|---|---|
| | Chr10 | | |
| rs3135819 | 121481608 | C | G |
| rs2912798 | 121481809 | G | A |
| rs3135814 | 121482512 | T | C |
| rs2288336 | 121487098 | G | A |
| rs3135797 | 121489543 | T | C |
| rs3135795 | 121489746 | G | T |
| rs12762633 | 121493574 | C | T |
| rs2912754 | 121502180 | C | T |
| rs3135772 | 121504102 | C | T |
| rs3135770 | 121504978 | C | A |
| rs7090018 | 121509375 | T | G |
| rs116941692 | 121510702 | C | T |
| rs78985527 | 121512025 | G | A |
| rs77295559 | 121526011 | C | T |
| rs17541509 | 121526106 | G | T |
| rs117061282 | 121530054 | A | G |
| rs11199993 | 121531750 | G | C |
| rs74985544 | 121532069 | G | A |
| rs3135752 | 121536644 | G | A |
| rs2912770 | 121536805 | G | T |
| rs2981431 | 121550230 | C | T |
| rs755793 | 121551357 | A | G |
| rs17542209 | 121558412 | C | T |
| rs56226109 | 121565644 | G | A |
| rs2912787 | 121566024 | T | C |
| rs3135730 | 121566280 | T | C |
| rs36048667 | 121569015 | C | T |
| rs3750817 | 121573063 | C | T |
| rs2981579 | 121577821 | A | G |
| rs17542768 | 121578300 | A | G |
| rs45631549 | 121578958 | C | T |
| rs45631614 | 121580813 | G | C |
| rs45631630 | 121581310 | G | A |
| rs2981575 | 121586602 | G | A |
| rs1219648 | 121586676 | A | G |

Continued on next page

**Supplementary Table S1**: SNPs included in Phase 1 (Continued)

| rsid | Position | Reference allele | Alternate allele |
|---|---|---|---|
| rs17542858 | 121589916 | G | A |
| rs2981582 | 121592803 | A | G |
| rs45631558 | 121597696 | C | T |
| rs41301031 | 121600213 | G | A |
| rs73356565 | 121606210 | C | T |
| rs74886369 | 121608587 | C | T |
| rs117813079 | 121612410 | G | A |
| rs10788190 | 121627515 | A | G |
| rs1631281 | 121631271 | T | G |
| rs7079914 | 121633995 | G | C |
| rs1696846 | 121638282 | G | C |
| rs7068354 | 121651426 | A | C |
| rs118181559 | 121658722 | G | T |
| rs117891398 | 121660849 | G | A |
| rs116850002 | 121661763 | G | A |
| rs12242201 | 121663344 | C | T |
| rs61874138 | 121666102 | T | C |
| rs1696837 | 121668128 | A | G |
| rs61874140 | 121676468 | A | G |
| rs1219487 | 121680765 | T | C |
| | Chr11 | | |
| rs10160464 | 69419318 | A | G |
| rs4980785 | 69419726 | C | T |
| rs7105934 | 69424973 | G | A |
| rs6606643 | 69427039 | G | A |
| rs7927237 | 69431044 | A | C |
| rs79373485 | 69432853 | C | T |
| rs61881566 | 69440357 | T | C |
| rs117796657 | 69441137 | C | A |
| rs74512832 | 69445812 | C | T |
| rs7394746 | 69447660 | T | C |
| rs7113550 | 69455000 | G | A |
| rs10501399 | 69455090 | C | T |
| rs4444099 | 69458108 | C | T |
| rs57217362 | 69463122 | C | T |
| rs10737153 | 69467061 | A | C |
| rs57506806 | 69468656 | A | G |
| rs117752342 | 69475035 | C | T |
| rs76987532 | 69481119 | C | T |
| rs587230 | 69485003 | A | G |
| rs1122316 | 69485307 | G | A |

Continued on next page

**Supplementary Table S1**: SNPs included in Phase 1 (Continued)

| rsid | Position | Reference allele | Alternate allele |
|---|---|---|---|
| rs4980661 | 69491811 | G | A |
| rs2298764 | 69493156 | T | C |
| rs637185 | 69493255 | C | T |
| rs117222887 | 69494966 | G | A |
| rs665172 | 69497221 | C | T |
| rs592081 | 69499499 | T | C |
| rs11601399 | 69505694 | G | A |
| rs76809977 | 69508496 | C | T |
| rs598828 | 69512840 | C | G |
| rs559664 | 69517492 | A | G |
| rs657315 | 69517944 | C | T |
| rs498931 | 69524849 | A | G |
| rs71465432 | 69528387 | C | T |
| rs75296154 | 69529047 | C | T |
| rs72929545 | 69534620 | G | A |
| rs1463618 | 69536953 | G | T |
| rs183782062 | 69539114 | G | T |
| rs79442425 | 69562741 | C | T |
| rs117717143 | 69563926 | G | A |
| rs2046494 | 69564065 | T | C |
| rs111929748 | 69564519 | G | T |
| rs117490805 | 69572513 | G | A |
| rs2156513 | 69577504 | T | G |
| rs625237 | 69577536 | G | A |
| rs638385 | 69578184 | G | A |
| rs72932461 | 69578903 | G | A |
| rs7108090 | 69579623 | A | G |
| rs79241527 | 69581315 | C | T |
| rs77997559 | 69589495 | A | G |
| rs7122472 | 69600013 | T | C |
| rs72932500 | 69603668 | A | G |
| rs75277033 | 69604545 | T | A |
| rs74412847 | 69606421 | A | G |
| rs113417932 | 69607996 | C | T |
| rs77772082 | 69608555 | C | T |
| rs57162717 | 69609602 | A | T |
| rs66516223 | 69616860 | G | A |
| | Chr16 | | |
| rs78338509 | 52486414 | A | G |
| rs12447430 | 52487716 | T | C |
| rs45545931 | 52489728 | T | C |

Continued on next page

**Supplementary Table S1**: SNPs included in Phase 1 (Continued)

| rsid | Position | Reference allele | Alternate allele |
|---|---|---|---|
| rs7186907 | 52493491 | G | A |
| rs45577538 | 52494687 | G | T |
| rs9302555 | 52494935 | T | G |
| rs78592318 | 52495195 | T | C |
| rs4784220 | 52501898 | T | C |
| rs8046985 | 52502436 | G | A |
| rs45542333 | 52506283 | G | T |
| rs78268044 | 52507901 | C | T |
| rs45454402 | 52514240 | A | G |
| rs45512493 | 52525814 | A | G |
| rs45587233 | 52530114 | T | A |
| rs45625634 | 52530262 | T | C |
| rs45577136 | 52532308 | A | G |
| rs45496893 | 52534218 | C | T |
| rs34750829 | 52535625 | G | A |
| rs9931232 | 52538920 | G | A |
| rs3803662 | 52552429 | A | G |
| rs45477396 | 52555435 | T | C |
| rs45493397 | 52557807 | C | T |
| rs75571494 | 52562272 | G | A |
| rs4594251 | 52563932 | C | T |
| rs4784227 | 52565276 | C | T |
| rs76543857 | 52569948 | C | T |
| rs11864809 | 52570221 | T | C |
| rs118162666 | 52572969 | G | A |
| rs9708611 | 52578753 | G | A |
| rs3104767 | 52590826 | G | T |
| rs45570038 | 52598777 | A | C |
| rs3112612 | 52601252 | G | A |
| rs45568636 | 52601401 | C | T |
| rs45584434 | 52603296 | G | A |
| rs45560737 | 52603720 | C | A |
| rs45575339 | 52604455 | C | T |
| rs45587544 | 52606294 | G | A |
| rs3112606 | 52616997 | T | C |
| rs11647405 | 52618868 | T | C |
| rs11075562 | 52619068 | G | A |
| rs3104805 | 52620954 | C | T |
| rs3112601 | 52621374 | C | G |
| rs117254441 | 52623216 | C | T |
| rs3112599 | 52623621 | A | G |

Continued on next page

**Supplementary Table S1**: SNPs included in Phase 1 (Continued)

| rsid | Position | Reference allele | Alternate allele |
|---|---|---|---|
| rs117389216 | 52624426 | T | C |
| rs56117730 | 52625523 | G | C |
| rs78841172 | 52626191 | T | C |
| rs76000465 | 52626967 | T | C |
| rs10521266 | 52627132 | G | C |
| rs3112596 | 52629063 | T | C |
| rs7188201 | 52632690 | A | G |
| rs111925335 | 52633030 | A | C |
| rs75127968 | 52634678 | G | A |
| rs3104816 | 52637858 | C | T |
| rs3104819 | 52641501 | A | G |
| rs3104824 | 52645181 | A | C |

Supplementary Table S2 shows the total number of unique patterns and the number of closed patterns discovered at each iteration of Phase 1. Iteration 0 corresponds to the initiation step; only the number of filtered patterns is given. Closed patterns are the patterns that last appear in a given iteration. A final untabulated iteration is carried out to ensure that the final pattern(s) disappear upon overlap (no pattern is overlapped with itself). The results are shown graphically in Supplementary Fig. S1.

**Supplementary Table S2**: Closed-pattern discovery, Phase 1

| Iteration | Total | Closed |
|---|---|---|
| | Chr10 | |
| 0 | 85 | 17 |
| 1 | 1342 | 44 |
| 2 | 29522 | 89 |
| 3 | 229571 | 186 |
| 4 | 251524 | 425 |
| 5 | 251099 | 959 |
| 6 | 250140 | 2017 |
| 7 | 248123 | 3914 |
| 8 | 244209 | 6996 |
| 9 | 237213 | 11428 |
| 10 | 225785 | 16912 |
| 11 | 208873 | 22547 |
| 12 | 186326 | 27097 |
| 13 | 159229 | 29605 |
| 14 | 129624 | 29789 |
| 15 | 99835 | 27850 |
| 16 | 71985 | 24106 |
| 17 | 47879 | 19008 |

Continued on next page

**Supplementary Table S2**: Closed-pattern discovery, Phase 1 (Continued)

| Iteration | Total | Closed |
|---|---|---|
| 18 | 28871 | 13360 |
| 19 | 15511 | 8199 |
| 20 | 7312 | 4323 |
| 21 | 2989 | 1940 |
| 22 | 1049 | 739 |
| 23 | 310 | 237 |
| 24 | 73 | 61 |
| 25 | 12 | 11 |
| 26 | 1 | 1 |
| | Chr11 | |
| 0 | 115 | 27 |
| 1 | 2898 | 77 |
| 2 | 75361 | 160 |
| 3 | 283854 | 300 |
| 4 | 285829 | 538 |
| 5 | 285291 | 940 |
| 6 | 284351 | 1570 |
| 7 | 282781 | 2462 |
| 8 | 280319 | 3616 |
| 9 | 276703 | 5076 |
| 10 | 271627 | 6933 |
| 11 | 264694 | 9373 |
| 12 | 255321 | 12551 |
| 13 | 242770 | 16399 |
| 14 | 226371 | 20497 |
| 15 | 205874 | 24156 |
| 16 | 181718 | 26689 |
| 17 | 155029 | 27675 |
| 18 | 127354 | 27055 |
| 19 | 100299 | 25017 |
| 20 | 75282 | 21849 |
| 21 | 53433 | 17915 |
| 22 | 35518 | 13667 |
| 23 | 21851 | 9589 |
| 24 | 12262 | 6097 |
| 25 | 6165 | 3449 |
| 26 | 2716 | 1697 |
| 27 | 1019 | 706 |
| 28 | 313 | 239 |
| 29 | 74 | 62 |
| 30 | 12 | 11 |

Continued on next page

**Supplementary Table S2**: Closed-pattern discovery, Phase 1 (Continued)

| Iteration | Total | Closed |
|---|---|---|
| 31 | 1 | 1 |
| | Chr16 | |
| 0 | 245 | 51 |
| 1 | 8444 | 114 |
| 2 | 146411 | 235 |
| 3 | 269142 | 455 |
| 4 | 268795 | 848 |
| 5 | 267947 | 1515 |
| 6 | 266432 | 2542 |
| 7 | 263890 | 4035 |
| 8 | 259855 | 6102 |
| 9 | 253753 | 8759 |
| 10 | 244994 | 11911 |
| 11 | 233083 | 15461 |
| 12 | 217622 | 19233 |
| 13 | 198389 | 22854 |
| 14 | 175535 | 25755 |
| 15 | 149780 | 27318 |
| 16 | 122462 | 27149 |
| 17 | 95313 | 25255 |
| 18 | 70058 | 21949 |
| 19 | 48109 | 17677 |
| 20 | 30432 | 12994 |
| 21 | 17438 | 8559 |
| 22 | 8879 | 4956 |
| 23 | 3923 | 2467 |
| 24 | 1456 | 1023 |
| 25 | 433 | 337 |
| 26 | 96 | 82 |
| 27 | 14 | 13 |
| 28 | 1 | 1 |

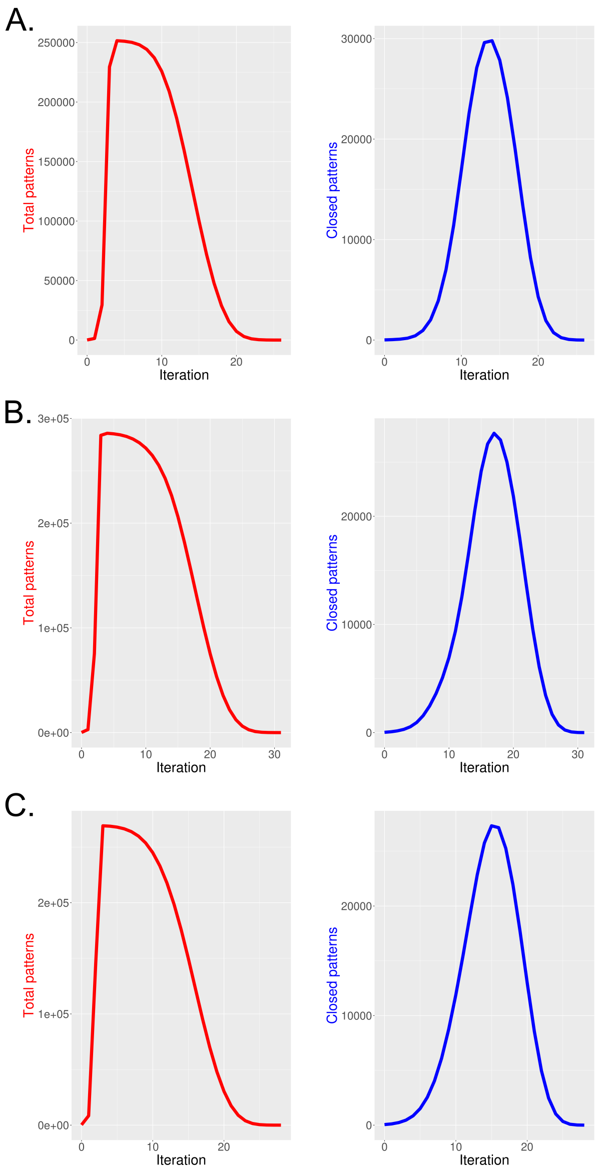

**Supplementary Figure S1:** The total (left) and closed (right) patterns found at each iteration of Phase 1 at 10q26 (chr10) (A), 11q13 (chr11) (B), and 16q12 (chr16) (C).

Supplementary Table S3 gives the SNPs selected in each region for Phase 2. All coordinates are GRCh38. Bolded h1 SNPs are shown for reference but were not included in Phase 2.

**Supplementary Table S3**: SNPs included in Phase 2

| rsid | Position | Reference allele | Alternate allele |
|---|---|---|---|
| | Chr10 | | |
| rs4752498 | 121065946 | C | T |
| rs7078330 | 121074650 | C | T |
| rs76468001 | 121075347 | C | T |
| rs117641945 | 121075645 | T | G |
| rs78493319 | 121082203 | C | T |
| rs118104738 | 121088861 | C | T |
| rs2130789 | 121092915 | T | C |
| rs72826685 | 121095693 | T | C |
| rs10886849 | 121098449 | C | T |
| rs2244506 | 121102856 | T | G |
| rs185196870 | 121108592 | C | A |
| rs10886852 | 121109082 | C | T |
| rs7907754 | 121110902 | A | G |
| rs72828403 | 121111614 | A | G |
| rs2254069 | 121116075 | G | A |
| rs80246243 | 121126040 | T | C |
| rs4436487 | 121126726 | T | C |
| rs78250596 | 121129666 | G | A |
| rs77170423 | 121130744 | T | C |
| rs12259125 | 121132720 | G | A |
| rs12777051 | 121136555 | C | A |
| rs17101493 | 121136678 | T | C |
| rs2901286 | 121141109 | C | A |
| rs80274247 | 121141924 | T | G |
| rs61874880 | 121152639 | G | A |
| rs11199800 | 121152876 | G | A |
| rs72631105 | 121155831 | G | A |
| rs11199805 | 121159692 | C | T |
| rs71484661 | 121170207 | C | T |
| rs56297542 | 121174596 | C | T |
| rs1907282 | 121182017 | G | A |
| rs11597363 | 121188377 | C | G |
| rs7089185 | 121189319 | G | A |
| rs11199825 | 121201341 | G | A |
| rs10788149 | 121207656 | G | A |
| rs12777794 | 121210296 | C | T |

Continued on next page

**Supplementary Table S3**: SNPs included in Phase 2 (Continued)

| rsid | Position | Reference allele | Alternate allele |
|---|---|---|---|
| rs12221275 | 121212884 | C | A |
| rs76847969 | 121215334 | T | C |
| rs75273759 | 121215359 | C | T |
| rs80351942 | 121217371 | G | T |
| rs1907227 | 121217582 | T | G |
| rs55740531 | 121223232 | T | C |
| rs76105384 | 121223582 | A | G |
| rs10886873 | 121225604 | C | T |
| rs72830621 | 121236753 | G | A |
| rs72830624 | 121241047 | A | G |
| rs116967076 | 121244065 | G | A |
| rs7899480 | 121244121 | C | T |
| rs11199860 | 121261519 | A | C |
| rs7076500 | 121262217 | G | A |
| rs4752526 | 121263729 | C | T |
| rs7915008 | 121265711 | A | G |
| rs12253779 | 121267364 | A | G |
| rs11199874 | 121273005 | G | A |
| rs78331385 | 121273943 | C | T |
| rs10788160 | 121274035 | G | A |
| rs80091641 | 121276743 | C | A |
| rs77914714 | 121286643 | T | G |
| rs12570783 | 121300385 | A | G |
| rs72830660 | 121300461 | C | T |
| rs75707486 | 121318187 | G | T |
| rs4752530 | 121321935 | G | T |
| rs117666755 | 121322790 | G | A |
| rs76866916 | 121329079 | G | A |
| rs4752536 | 121333293 | A | G |
| rs116988595 | 121334710 | T | A |
| rs35098964 | 121335498 | C | T |
| rs10466213 | 121336954 | C | G |
| rs9421411 | 121337045 | G | A |
| rs10466215 | 121337233 | T | C |
| rs146809981 | 121347140 | A | G |
| rs7893305 | 121350085 | A | G |
| rs12256422 | 121350961 | G | A |
| rs80126005 | 121355935 | G | A |
| rs7071430 | 121356173 | C | T |
| rs2420932 | 121358704 | T | C |
| rs60348293 | 121359423 | G | A |

Continued on next page

**Supplementary Table S3**: SNPs included in Phase 2 (Continued)

| rsid | Position | Reference allele | Alternate allele |
|---|---|---|---|
| rs17601696 | 121360522 | C | T |
| rs72830688 | 121361996 | G | A |
| rs72830689 | 121368352 | G | C |
| rs12220114 | 121370981 | A | G |
| rs11199936 | 121374968 | C | G |
| rs10788173 | 121375686 | C | T |
| rs10886929 | 121376210 | T | C |
| rs75546829 | 121377689 | C | T |
| rs1896403 | 121378757 | T | A |
| rs73362352 | 121383742 | G | A |
| rs4752546 | 121389242 | A | G |
| rs1896405 | 121391808 | A | G |
| rs76834389 | 121391820 | C | T |
| rs4752550 | 121391974 | C | T |
| rs74158584 | 121406190 | A | G |
| rs2162541 | 121408577 | C | T |
| rs12413779 | 121410999 | T | G |
| rs74158588 | 121412857 | C | G |
| rs76352137 | 121415243 | C | A |
| rs10788176 | 121420794 | T | C |
| rs12766813 | 121423158 | T | C |
| rs11199954 | 121426042 | G | A |
| rs17603569 | 121426300 | A | G |
| rs11199957 | 121428653 | C | T |
| rs17101979 | 121430598 | T | G |
| rs11199964 | 121434484 | T | C |
| **rs3135795** | **121489746** | **G** | **T** |
| **rs116941692** | **121510702** | **C** | **T** |
| **rs755793** | **121551357** | **A** | **G** |
| **rs56226109** | **121565644** | **G** | **A** |
| **rs2981579** | **121577821** | **A** | **G** |
| **rs45631549** | **121578958** | **C** | **T** |
| **rs45631614** | **121580813** | **G** | **C** |
| **rs45631630** | **121581310** | **G** | **A** |
| **rs17542858** | **121589916** | **G** | **A** |
| **rs118181559** | **121658722** | **G** | **T** |
| | Chr11 | | |
| rs2376558 | 69083946 | T | C |
| rs896973 | 69083998 | G | T |
| rs61881030 | 69085616 | C | A |
| rs149949063 | 69085685 | A | G |

Continued on next page

**Supplementary Table S3**: SNPs included in Phase 2 (Continued)

| rsid | Position | Reference allele | Alternate allele |
|---|---|---|---|
| rs144905179 | 69085729 | G | A |
| rs1123665 | 69087729 | T | G |
| rs3829241 | 69087895 | G | A |
| rs74674490 | 69092678 | A | G |
| rs72930627 | 69099653 | C | A |
| rs72930631 | 69100367 | T | C |
| rs11228502 | 69110120 | T | C |
| rs12577213 | 69114683 | C | A |
| rs73520512 | 69114776 | C | T |
| rs3892895 | 69117287 | A | G |
| rs10896431 | 69119549 | C | T |
| rs7116054 | 69124227 | C | T |
| rs74897859 | 69126381 | C | T |
| rs17308715 | 69127338 | C | T |
| rs67039008 | 69130541 | G | A |
| rs55703863 | 69133728 | G | C |
| rs17149775 | 69142111 | A | G |
| rs117236867 | 69145367 | A | G |
| rs11228530 | 69151517 | A | C |
| rs17309046 | 69151962 | T | C |
| rs56134379 | 69153340 | G | A |
| rs72932540 | 69154575 | A | G |
| rs12418948 | 69159054 | C | T |
| rs78033785 | 69168018 | C | T |
| rs74342245 | 69171308 | G | A |
| rs35435383 | 69180342 | G | A |
| rs4495899 | 69191192 | T | G |
| rs117694794 | 69195706 | C | T |
| rs10792029 | 69199415 | A | G |
| rs10896445 | 69200174 | T | C |
| rs78208050 | 69201946 | A | G |
| rs61881109 | 69209346 | G | A |
| rs75590802 | 69210118 | G | A |
| rs11228565 | 69211113 | G | A |
| rs7937094 | 69212239 | C | T |
| rs7931342 | 69227030 | T | G |
| rs10896449 | 69227200 | A | G |
| rs7130881 | 69228491 | A | G |
| rs7119988 | 69248404 | G | A |
| rs111762835 | 69250616 | G | A |
| rs4930672 | 69251764 | G | A |

Continued on next page

**Supplementary Table S3**: SNPs included in Phase 2 (Continued)

| rsid | Position | Reference allele | Alternate allele |
|---|---|---|---|
| rs11228599 | 69258819 | G | T |
| rs7940107 | 69260303 | G | A |
| rs72930267 | 69263523 | A | T |
| rs7946255 | 69263730 | T | C |
| rs116951063 | 69265264 | C | T |
| rs118004919 | 69267799 | G | A |
| rs35637432 | 69269130 | G | A |
| rs34737133 | 69275141 | C | T |
| rs11825015 | 69277004 | C | T |
| rs78656034 | 69285720 | C | T |
| rs74551015 | 69286582 | G | A |
| rs117821797 | 69287175 | C | T |
| rs12275849 | 69287948 | T | C |
| rs56103266 | 69289127 | G | C |
| rs117131950 | 69295686 | G | A |
| rs7103126 | 69295926 | T | C |
| rs116926312 | 69296009 | C | T |
| rs11539762 | 69296043 | G | A |
| rs142581206 | 69296259 | G | T |
| rs11228610 | 69296261 | G | C |
| rs12274095 | 69296300 | C | A |
| rs73512137 | 69296517 | C | T |
| rs11601693 | 69302071 | G | A |
| rs10896461 | 69306180 | A | G |
| rs12224376 | 69306623 | G | A |
| rs78919175 | 69309047 | C | T |
| rs7124547 | 69309919 | C | T |
| rs11603814 | 69311104 | C | G |
| rs72932105 | 69311184 | C | T |
| rs12789955 | 69311667 | T | G |
| rs7481709 | 69317083 | T | C |
| rs115223540 | 69321285 | C | T |
| rs28378931 | 69321751 | A | G |
| rs7102705 | 69328516 | A | G |
| rs117186144 | 69338687 | C | T |
| rs79487139 | 69352233 | C | T |
| rs11600497 | 69358550 | C | A |
| rs72932198 | 69360358 | G | A |
| rs12802601 | 69361135 | T | C |
| rs12796465 | 69367952 | A | G |
| rs4275647 | 69371038 | C | T |

Continued on next page

**Supplementary Table S3**: SNPs included in Phase 2 (Continued)

| rsid | Position | Reference allele | Alternate allele |
|---|---|---|---|
| rs61882193 | 69377704 | A | G |
| rs12790064 | 69382816 | A | G |
| rs11263638 | 69386345 | G | A |
| rs11263641 | 69392822 | T | C |
| rs76855139 | 69409182 | C | T |
| rs74471298 | 69414699 | G | A |
| **rs7105934** | **69424973** | **G** | **A** |
| **rs79373485** | **69432853** | **C** | **T** |
| **rs4444099** | **69458108** | **C** | **T** |
| **rs117752342** | **69475035** | **C** | **T** |
| **rs1122316** | **69485307** | **G** | **A** |
| **rs76809977** | **69508496** | **C** | **T** |
| **rs559664** | **69517492** | **A** | **G** |
| **rs657315** | **69517944** | **C** | **T** |
| **rs498931** | **69524849** | **A** | **G** |
| **rs71465432** | **69528387** | **C** | **T** |
| **rs183782062** | **69539114** | **G** | **T** |
| **rs79442425** | **69562741** | **C** | **T** |
| **rs111929748** | **69564519** | **G** | **T** |
| **rs117490805** | **69572513** | **G** | **A** |
| **rs79241527** | **69581315** | **C** | **T** |
| **rs72932500** | **69603668** | **A** | **G** |
| **rs57162717** | **69609602** | **A** | **T** |
| | Chr16 | | |
| rs1420281 | 52168409 | T | C |
| rs60526343 | 52168958 | T | G |
| rs72792181 | 52170386 | G | C |
| rs9939929 | 52170892 | T | C |
| rs3095540 | 52177742 | C | T |
| rs11649329 | 52178212 | C | T |
| rs116988362 | 52184126 | T | C |
| rs4785118 | 52185522 | G | A |
| rs17268400 | 52186834 | T | C |
| rs117108822 | 52198541 | C | A |
| rs72792197 | 52205173 | A | C |
| rs117285516 | 52207069 | C | A |
| rs116048640 | 52210614 | C | A |
| rs7205520 | 52212173 | A | G |
| rs118147607 | 52213044 | C | T |
| rs113755361 | 52214851 | C | A |
| rs113083456 | 52223616 | C | T |

Continued on next page

**Supplementary Table S3**: SNPs included in Phase 2 (Continued)

| rsid | Position | Reference allele | Alternate allele |
|---|---|---|---|
| rs12930272 | 52231880 | G | A |
| rs1345316 | 52246778 | G | A |
| rs2287071 | 52254786 | A | G |
| rs1362376 | 52258027 | T | C |
| rs2540701 | 52264270 | G | A |
| rs112797109 | 52280250 | C | T |
| rs72794126 | 52284113 | C | T |
| rs74393050 | 52289258 | G | A |
| rs17270950 | 52289574 | T | C |
| rs117142114 | 52294754 | T | C |
| rs117832727 | 52307932 | G | C |
| rs9938443 | 52310370 | G | A |
| rs4784165 | 52313907 | T | G |
| **rs78338509** | **52486414** | **A** | **G** |
| **rs12447430** | **52487716** | **T** | **C** |
| **rs45577538** | **52494687** | **G** | **T** |
| **rs8046985** | **52502436** | **G** | **A** |
| **rs45542333** | **52506283** | **G** | **T** |
| **rs78268044** | **52507901** | **C** | **T** |
| **rs45454402** | **52514240** | **A** | **G** |
| **rs45512493** | **52525814** | **A** | **G** |
| **rs34750829** | **52535625** | **G** | **A** |
| **rs9931232** | **52538920** | **G** | **A** |
| **rs4594251** | **52563932** | **C** | **T** |
| **rs4784227** | **52565276** | **C** | **T** |
| **rs45584434** | **52603296** | **G** | **A** |
| **rs45560737** | **52603720** | **C** | **A** |
| **rs45575339** | **52604455** | **C** | **T** |
| **rs45587544** | **52606294** | **G** | **A** |
| **rs78841172** | **52626191** | **T** | **C** |
| **rs76000465** | **52626967** | **T** | **C** |
| **rs111925335** | **52633030** | **A** | **C** |
| **rs75127968** | **52634678** | **G** | **A** |
| rs79291013 | 52794506 | C | G |
| rs8044632 | 52797054 | T | C |
| rs117746278 | 52812434 | G | A |
| rs9939896 | 52817953 | C | T |
| rs2024446 | 52819595 | G | A |
| rs17298178 | 52820762 | T | C |
| rs79993873 | 52827626 | A | C |
| rs78320298 | 52829163 | A | G |

Continued on next page

**Supplementary Table S3**: SNPs included in Phase 2 (Continued)

| rsid | Position | Reference allele | Alternate allele |
|---|---|---|---|
| rs62041381 | 52831633 | A | C |
| rs117222080 | 52832479 | G | T |
| rs117551222 | 52834000 | T | C |
| rs7500472 | 52835202 | G | A |
| rs72810045 | 52835238 | G | A |
| rs12446016 | 52841596 | T | G |
| rs73591253 | 52856204 | C | T |
| rs150704739 | 52861748 | C | T |
| rs113964117 | 52863621 | G | A |
| rs1345327 | 52863982 | T | C |
| rs142367768 | 52869028 | C | T |
| rs4783791 | 52870135 | T | G |
| rs117911537 | 52877564 | C | T |
| rs1344484 | 52878387 | T | C |
| rs16951771 | 52884471 | G | A |
| rs73597288 | 52887544 | G | A |
| rs12919591 | 52889126 | A | G |
| rs9630629 | 52906807 | A | C |
| rs6499147 | 52909787 | T | C |
| rs12922473 | 52910315 | A | G |
| rs78322982 | 52912824 | G | A |
| rs6499148 | 52916696 | C | A |
| rs62043282 | 52917069 | C | A |
| rs113717347 | 52923006 | G | A |
| rs62043323 | 52925609 | G | A |
| rs12924810 | 52929042 | C | A |
| rs80356178 | 52929430 | G | A |
| rs34261756 | 52929642 | T | C |
| rs117400881 | 52932025 | T | C |
| rs1477029 | 52932261 | G | A |
| rs7199322 | 52941691 | T | G |
| rs78831406 | 52942873 | C | T |
| rs4784276 | 52950865 | T | C |
| rs77419902 | 52960811 | C | T |
| rs78724580 | 52960991 | G | A |
| rs72812119 | 52962135 | A | G |
| rs72812122 | 52963810 | G | A |
| rs62047145 | 52966065 | C | T |
| rs76365437 | 52966273 | G | A |
| rs13335861 | 52982252 | A | G |
| rs4352043 | 52983666 | T | C |

Continued on next page

**Supplementary Table S3**: SNPs included in Phase 2 (Continued)

| rsid | Position | Reference allele | Alternate allele |
|---|---|---|---|
| rs36099013 | 52986896 | G | A |
| rs72812136 | 52988181 | C | T |
| rs79815180 | 52992846 | C | T |
| rs74018575 | 53001419 | A | C |
| rs72812140 | 53002388 | G | T |
| rs34382563 | 53003215 | C | T |
| rs12598778 | 53006686 | T | C |
| rs56085964 | 53010795 | T | C |
| rs7499206 | 53012222 | T | C |
| rs12922187 | 53019595 | A | G |
| rs8051957 | 53020498 | A | G |
| rs8050297 | 53020579 | G | A |
| rs79721160 | 53021625 | G | A |
| rs79088862 | 53022198 | G | A |
| rs4784287 | 53023458 | C | T |
| rs117297253 | 53023797 | G | A |
| rs72797618 | 53030122 | T | C |
| rs9933155 | 53033775 | C | T |
| rs56278234 | 53034901 | C | T |
| rs3931698 | 53036913 | G | T |
| rs62049054 | 53042219 | T | C |
| rs113465432 | 53051819 | C | T |
| rs11641638 | 53053996 | G | A |

Supplementary Table S4 shows the total number of unique patterns and the number of closed patterns discovered at each iteration of Phase 2. Iteration 0 corresponds to the initiation step; only the number of filtered patterns is given. Closed patterns are the patterns that last appear in a given iteration. A final untabulated iteration is carried out to ensure that the final pattern(s) disappear upon overlap (no pattern is overlapped with itself).

**Supplementary Table S4**: Closed-pattern discovery, Phase 2

| Iteration | Total | Closed |
|---|---|---|
| | Chr10 | |
| 0 | 39 | 27 |
| 1 | 670 | 222 |
| 2 | 29748 | 789 |
| 3 | 339253 | 1855 |
| 4 | 394926 | 3427 |
| 5 | 391499 | 5518 |
| 6 | 385981 | 8091 |
| 7 | 377890 | 10919 |

Continued on next page

**Supplementary Table S4**: Closed-pattern discovery, Phase 2 (Continued)

| Iteration | Total | Closed |
|---|---|---|
| 8 | 366971 | 13810 |
| 9 | 353161 | 16670 |
| 10 | 336491 | 19365 |
| 11 | 317126 | 21759 |
| 12 | 295367 | 23871 |
| 13 | 271496 | 25827 |
| 14 | 245669 | 27581 |
| 15 | 218088 | 28825 |
| 16 | 189263 | 29220 |
| 17 | 160043 | 28600 |
| 18 | 131443 | 26982 |
| 19 | 104461 | 24500 |
| 20 | 79961 | 21360 |
| 21 | 58601 | 17795 |
| 22 | 40806 | 14047 |
| 23 | 26759 | 10392 |
| 24 | 16367 | 7131 |
| 25 | 9236 | 4495 |
| 26 | 4741 | 2571 |
| 27 | 2170 | 1310 |
| 28 | 860 | 578 |
| 29 | 282 | 211 |
| 30 | 71 | 59 |
| 31 | 12 | 11 |
| 32 | 1 | 1 |
| | Chr11 | |
| 0 | 28 | 21 |
| 1 | 349 | 142 |
| 2 | 11196 | 554 |
| 3 | 143202 | 1574 |
| 4 | 165707 | 3572 |
| 5 | 162135 | 6742 |
| 6 | 155393 | 10835 |
| 7 | 144558 | 15151 |
| 8 | 129407 | 18737 |
| 9 | 110670 | 20757 |
| 10 | 89913 | 20838 |
| 11 | 69075 | 19089 |
| 12 | 49986 | 15981 |
| 13 | 34005 | 12218 |
| 14 | 21787 | 8530 |

Continued on next page

**Supplementary Table S4**: Closed-pattern discovery, Phase 2 (Continued)

| Iteration | Total | Closed |
|---|---|---|
| 15 | 13257 | 5475 |
| 16 | 7782 | 3305 |
| 17 | 4477 | 1953 |
| 18 | 2524 | 1175 |
| 19 | 1349 | 711 |
| 20 | 638 | 394 |
| 21 | 244 | 176 |
| 22 | 68 | 56 |
| 23 | 12 | 11 |
| 24 | 1 | 1 |
| | Chr16 | |
| 0 | 25 | 23 |
| 1 | 295 | 237 |
| 2 | 12076 | 1346 |
| 3 | 278896 | 4807 |
| 4 | 362193 | 12080 |
| 5 | 350113 | 23123 |
| 6 | 326990 | 35523 |
| 7 | 291467 | 45646 |
| 8 | 245821 | 50690 |
| 9 | 195131 | 49801 |
| 10 | 145330 | 43958 |
| 11 | 101372 | 35192 |
| 12 | 66180 | 25719 |
| 13 | 40461 | 17266 |
| 14 | 23195 | 10719 |
| 15 | 12476 | 6194 |
| 16 | 6282 | 3345 |
| 17 | 2937 | 1681 |
| 18 | 1256 | 775 |
| 19 | 481 | 321 |
| 20 | 160 | 116 |
| 21 | 44 | 35 |
| 22 | 9 | 8 |
| 23 | 1 | 1 |

#### 5. Haplotype details

Supplementary Table S5 gives the SNP-alleles of h1 (reduced) the full-length haplotypes identified in Phase 2. As all haplotypes are subtypes of h1, the position of h1 in the haplotype is indicated.

**Supplementary Table S5**: Haplotype alleles

| Variable | SNP[allele] |
|---|---|
| Chr10 | |
| h1 | rs3135795[G], rs116941692[C], rs755793[A], rs56226109[G], rs2981579[A], rs45631549[C], rs45631614[G], rs45631630[G], rs17542858[G], rs118181559[G] |
| h2 | rs4752498[T], rs7078330[C], rs76468001[C], rs117641945[T], rs78493319[C], rs118104738[C], rs2130789[T], rs72826685[T], rs10886849[C], rs2244506[T], rs185196870[C], rs10886852[C], rs7907754[A], rs72828403[A], rs2254069[G], rs80246243[T], rs4436487[T], rs78250596[G], rs77170423[T], rs12259125[G], rs12777051[C], rs61874880[G], rs11199800[G], rs72631105[G], rs71484661[C], rs1907282[G], rs11597363[C], rs7089185[G], rs10788149[A], rs12777794[C], rs76847969[T], rs75273759[C], rs80351942[G], rs55740531[T], rs76105384[A], rs72830624[A], rs116967076[G], rs4752526[T], rs12253779[A], rs78331385[C], rs80091641[C], rs77914714[T], rs72830660[C], rs75707486[G], rs117666755[G], rs116988595[T], rs146809981[A], rs12256422[G], rs80126005[G], rs7071430[T], rs2420932[T], rs17601696[C], rs72830688[G], rs72830689[G], rs12220114[A], rs11199936[C], rs10788173[C], rs75546829[C], rs1896403[A], rs1896405[G], rs76834389[C], rs74158584[A], rs12413779[T], rs74158588[C], rs76352137[C], rs12766813[T], rs17603569[A], rs17101979[T], rs11199964[C], h1 |

Continued on next page

**Supplementary Table S5**: Haplotype alleles (Continued)

| Variable | SNP[allele] |
|---|---|
| h3 | rs4752498[T], rs7078330[C], rs76468001[C], rs117641945[T], rs78493319[C], rs118104738[C], rs2130789[T], rs72826685[T], rs10886849[C], rs2244506[T], rs185196870[C], rs7907754[A], rs72828403[A], rs2254069[G], rs80246243[T], rs4436487[T], rs78250596[G], rs77170423[T], rs12259125[G], rs12777051[C], rs17101493[T], rs2901286[C], rs80274247[T], rs61874880[G], rs11199805[C], rs71484661[C], rs56297542[C], rs11597363[C], rs11199825[G], rs12777794[C], rs12221275[C], rs76847969[T], rs75273759[C], rs80351942[G], rs55740531[T], rs76105384[A], rs72830621[G], rs72830624[A], rs116967076[G], rs4752526[T], rs12253779[A], rs78331385[C], rs80091641[C], rs77914714[T], rs72830660[C], rs75707486[G], rs117666755[G], rs76866916[G], rs4752536[G], rs116988595[T], rs10466213[G], rs9421411[G], rs146809981[A], rs7893305[A], rs12256422[G], rs80126005[G], rs7071430[T], rs2420932[T], rs60348293[G], rs17601696[C], rs72830688[G], rs72830689[G], rs12220114[A], rs11199936[C], rs10788173[C], rs75546829[C], rs1896403[A], rs73362352[G], rs4752546[A], rs76834389[C], rs74158584[A], rs2162541[T], rs12766813[C], rs11199954[G], rs17603569[A], rs11199957[C], rs17101979[T], h1 |

Continued on next page

**Supplementary Table S5**: Haplotype alleles (Continued)

| Variable | SNP[allele] |
| --- | --- |
| h4 | rs7078330[C], rs76468001[C], rs117641945[T], rs78493319[C], rs118104738[C], rs2130789[T], rs10886849[C], rs2244506[T], rs185196870[C], rs10886852[C], rs7907754[A], rs72828403[A], rs2254069[G], rs80246243[T], rs4436487[T], rs77170423[T], rs12259125[G], rs12777051[C], rs17101493[T], rs2901286[C], rs80274247[T], rs61874880[G], rs72631105[G], rs11199805[C], rs71484661[C], rs1907282[A], rs7089185[A], rs11199825[G], rs12777794[C], rs12221275[C], rs76847969[T], rs75273759[C], rs1907227[T], rs55740531[T], rs76105384[A], rs72830621[G], rs72830624[A], rs7899480[C], rs11199860[A], rs4752526[T], rs12253779[A], rs78331385[C], rs80091641[C], rs77914714[T], rs75707486[G], rs4752530[G], rs117666755[G], rs76866916[G], rs4752536[A], rs116988595[T], rs35098964[C], rs10466215[T], rs146809981[A], rs12256422[G], rs80126005[G], rs60348293[G], rs17601696[C], rs72830688[G], rs72830689[G], rs11199936[C], rs10886929[T], rs75546829[C], rs1896403[A], rs73362352[G], rs4752546[A], rs1896405[G], rs4752550[T], rs74158584[A], rs12413779[T], rs76352137[C], rs12766813[C], rs11199954[G], rs17603569[A], rs11199957[C], rs17101979[T], h1 |

Continued on next page

**Supplementary Table S5**: Haplotype alleles (Continued)

| Variable | SNP[allele] |
|---|---|
| h5 | rs4752498[T], rs7078330[C], rs76468001[C], rs117641945[T], rs78493319[C], rs118104738[C], rs2130789[T], rs72826685[T], rs10886849[C], rs2244506[T], rs185196870[C], rs7907754[A], rs72828403[A], rs2254069[G], rs80246243[T], rs4436487[T], rs78250596[G], rs77170423[T], rs12259125[G], rs12777051[C], rs17101493[T], rs2901286[C], rs80274247[T], rs61874880[G], rs11199800[A], rs11199805[C], rs71484661[C], rs56297542[C], rs11597363[C], rs11199825[G], rs12777794[C], rs12221275[C], rs76847969[T], rs75273759[C], rs80351942[G], rs1907227[T], rs55740531[T], rs76105384[A], rs10886873[T], rs72830624[A], rs116967076[G], rs7899480[C], rs11199860[A], rs7076500[A], rs4752526[T], rs12253779[A], rs78331385[C], rs80091641[C], rs77914714[T], rs12570783[A], rs75707486[G], rs117666755[G], rs76866916[G], rs4752536[A], rs116988595[T], rs35098964[C], rs10466215[T], rs146809981[A], rs7893305[A], rs12256422[G], rs80126005[G], rs7071430[C], rs2420932[T], rs60348293[G], rs17601696[C], rs12220114[G], rs11199936[C], rs10788173[C], rs75546829[C], rs1896403[A], rs73362352[G], rs76834389[C], rs4752550[C], rs74158584[A], rs2162541[C], rs12413779[T], rs74158588[C], rs76352137[C], rs11199954[G], rs17603569[A], rs11199957[C], rs17101979[T], rs11199964[T], h1 |

Continued on next page

**Supplementary Table S5**: Haplotype alleles (Continued)

| Variable | SNP[allele] |
|---|---|
| h6 | rs76468001[C], rs117641945[T], rs78493319[C], rs118104738[C], rs2130789[T], rs72826685[T], rs2244506[G], rs185196870[C], rs72828403[A], rs80246243[T], rs4436487[T], rs78250596[G], rs77170423[T], rs12259125[G], rs17101493[T], rs2901286[C], rs80274247[T], rs61874880[G], rs72631105[G], rs11199805[C], rs71484661[C], rs56297542[C], rs1907282[A], rs11597363[C], rs7089185[A], rs11199825[G], rs12777794[C], rs12221275[C], rs76847969[T], rs75273759[C], rs80351942[G], rs1907227[T], rs55740531[T], rs76105384[A], rs10886873[C], rs72830621[G], rs72830624[A], rs116967076[G], rs7899480[C], rs11199860[A], rs4752526[T], rs12253779[A], rs78331385[C], rs80091641[C], rs77914714[T], rs12570783[G], rs72830660[C], rs75707486[G], rs4752530[G], rs117666755[G], rs76866916[G], rs4752536[G], rs116988595[T], rs35098964[C], rs10466213[G], rs9421411[G], rs146809981[A], rs12256422[G], rs80126005[G], rs60348293[G], rs17601696[C], rs72830688[G], rs72830689[G], rs11199936[C], rs10788173[T], rs10886929[T], rs75546829[C], rs1896403[A], rs73362352[G], rs4752546[G], rs1896405[G], rs76834389[C], rs4752550[C], rs74158584[A], rs2162541[C], rs12413779[T], rs74158588[C], rs76352137[C], rs10788176[C], rs12766813[T], rs11199954[G], rs17603569[A], rs17101979[T], rs11199964[T], h1 |

Continued on next page

**Supplementary Table S5**: Haplotype alleles (Continued)

| Variable | SNP[allele] |
|---|---|
| h7 | rs4752498[T], rs7078330[C], rs76468001[C], rs117641945[T], rs78493319[C], rs118104738[C], rs2130789[T], rs72826685[T], rs10886849[C], rs2244506[T], rs185196870[C], rs10886852[C], rs7907754[A], rs72828403[A], rs2254069[G], rs80246243[T], rs4436487[T], rs78250596[G], rs77170423[T], rs12259125[G], rs12777051[C], rs17101493[T], rs2901286[C], rs80274247[T], rs61874880[G], rs11199800[G], rs11199805[C], rs56297542[C], rs1907282[A], rs11597363[C], rs7089185[A], rs11199825[G], rs10788149[G], rs12221275[C], rs76847969[T], rs75273759[C], rs80351942[G], rs1907227[T], rs55740531[T], rs76105384[A], rs72830621[G], rs72830624[A], rs116967076[G], rs7899480[C], rs11199860[A], rs7076500[A], rs4752526[T], rs7915008[G], rs12253779[A], rs11199874[G], rs78331385[C], rs10788160[G], rs80091641[C], rs77914714[T], rs12570783[G], rs72830660[C], rs75707486[G], rs4752530[G], rs117666755[G], rs76866916[G], rs4752536[G], rs116988595[T], rs35098964[C], rs10466213[G], rs9421411[G], rs10466215[C], rs146809981[A], rs12256422[G], rs80126005[G], rs60348293[G], rs17601696[C], rs72830688[G], rs72830689[G], rs11199936[C], rs10788173[T], rs10886929[T], rs75546829[C], rs1896403[A], rs73362352[G], rs1896405[G], rs76834389[C], rs74158584[A], rs2162541[C], rs12413779[T], rs74158588[C], rs76352137[C], rs11199954[G], rs17603569[A], rs11199957[C], rs17101979[T], rs11199964[T], h1 |
| Chr11 | |
| h1 | rs7105934[G], rs79373485[C], rs4444099[C], rs117752342[C], rs1122316[G], rs76809977[C], rs559664[G], rs657315[C], rs498931[A], rs71465432[C], rs183782062[G], rs79442425[C], rs111929748[G], rs117490805[G], rs79241527[C], rs72932500[A], rs57162717[A] |

Continued on next page

**Supplementary Table S5**: Haplotype alleles (Continued)

| Variable | SNP[allele] |
|---|---|
| h2 | rs2376558[C], rs149949063[A], rs144905179[G], rs74674490[A], rs72930627[C], rs72930631[T], rs11228502[T], rs12577213[C], rs73520512[C], rs10896431[C], rs74897859[C], rs55703863[G], rs17149775[A], rs117236867[A], rs11228530[A], rs17309046[T], rs56134379[G], rs72932540[A], rs78033785[C], rs74342245[G], rs35435383[G], rs117694794[C], rs10792029[G], rs78208050[A], rs61881109[G], rs75590802[G], rs7937094[C], rs7931342[T], rs10896449[A], rs7130881[A], rs7119988[G], rs111762835[G], rs4930672[G], rs11228599[G], rs7940107[G], rs72930267[A], rs7946255[T], rs116951063[C], rs118004919[G], rs35637432[G], rs34737133[C], rs11825015[C], rs78656034[C], rs117821797[C], rs56103266[G], rs117131950[G], rs7103126[C], rs116926312[C], rs12274095[C], rs10896461[G], rs78919175[C], rs7124547[T], rs12789955[T], rs115223540[C], rs28378931[A], rs117186144[C], rs72932198[G], rs12802601[T], rs12796465[A], rs61882193[A], rs12790064[G], rs11263638[G], rs74471298[G], h1 |
| h3 | rs2376558[C], rs61881030[C], rs149949063[A], rs144905179[G], rs74674490[A], rs12577213[C], rs73520512[C], rs3892895[G], rs7116054[C], rs74897859[C], rs17308715[C], rs67039008[A], rs17149775[G], rs117236867[A], rs11228530[A], rs17309046[T], rs56134379[G], rs72932540[A], rs12418948[C], rs78033785[C], rs74342245[G], rs35435383[G], rs117694794[C], rs10792029[G], rs78208050[A], rs61881109[G], rs75590802[G], rs7937094[C], rs7931342[G], rs10896449[G], rs7130881[A], rs111762835[G], rs4930672[G], rs116951063[C], rs118004919[G], rs35637432[G], rs34737133[C], rs78656034[C], rs74551015[G], rs117821797[C], rs12275849[T], rs117131950[G], rs7103126[T], rs11539762[G], rs142581206[G], rs11228610[G], rs12274095[C], rs73512137[C], rs12224376[G], rs11603814[C], rs72932105[C], rs12789955[T], rs117186144[C], rs79487139[C], rs11600497[C], rs72932198[G], rs12802601[T], rs12790064[G], rs76855139[C], rs74471298[G], h1 |

Continued on next page

**Supplementary Table S5**: Haplotype alleles (Continued)

| Variable | SNP[allele] |
|---|---|
| h4 | rs2376558[C], rs61881030[C], rs149949063[A], rs144905179[G], rs74674490[A], rs12577213[C], rs73520512[C], rs3892895[G], rs7116054[C], rs74897859[C], rs17308715[C], rs67039008[A], rs55703863[G], rs117236867[A], rs11228530[A], rs17309046[T], rs56134379[G], rs78033785[C], rs74342245[G], rs35435383[G], rs117694794[C], rs10792029[G], rs78208050[A], rs61881109[G], rs75590802[G], rs11228565[G], rs7937094[C], rs7130881[A], rs7119988[G], rs111762835[G], rs4930672[G], rs11228599[G], rs7940107[G], rs72930267[A], rs7946255[T], rs116951063[C], rs118004919[G], rs35637432[G], rs34737133[C], rs11825015[C], rs78656034[C], rs74551015[G], rs117821797[C], rs12275849[T], rs56103266[G], rs117131950[G], rs116926312[C], rs11539762[G], rs142581206[G], rs11228610[G], rs12274095[C], rs12224376[G], rs7124547[T], rs11603814[G], rs115223540[C], rs28378931[A], rs117186144[C], rs79487139[C], rs11600497[C], rs72932198[G], rs12802601[T], rs12796465[A], rs4275647[C], rs61882193[A], rs12790064[G], rs11263638[G], rs11263641[T], rs76855139[C], rs74471298[G], h1 |
| Chr16 | |
| h1 | rs78338509[A], rs12447430[T], rs45577538[G], rs8046985[A], rs45542333[G], rs78268044[C], rs45454402[A], rs45512493[A], rs34750829[G], rs9931232[A], rs4594251[C], rs4784227[T], rs45584434[G], rs45560737[C], rs45575339[C], rs45587544[G], rs78841172[T], rs76000465[T], rs111925335[A], rs75127968[G] |

Continued on next page

**Supplementary Table S5**: Haplotype alleles (Continued)

| Variable | SNP[allele] |
|---|---|
| h2 | rs1420281[T], rs60526343[T], rs72792181[G], rs3095540[T], rs11649329[C], rs116988362[T], rs4785118[G], rs17268400[T], rs117108822[C], rs72792197[A], rs117285516[C], rs116048640[C], rs7205520[A], rs118147607[C], rs113755361[C], rs113083456[C], rs1345316[A], rs2287071[A], rs2540701[G], rs112797109[C], rs72794126[C], rs17270950[T], rs117142114[T], rs117832727[G], rs9938443[A], h1, rs79291013[C], rs117746278[G], rs17298178[T], rs79993873[A], rs78320298[A], rs62041381[A], rs117222080[G], rs117551222[T], rs7500472[G], rs73591253[C], rs150704739[C], rs113964117[G], rs1345327[C], rs142367768[C], rs117911537[C], rs16951771[G], rs73597288[G], rs12919591[A], rs12922473[A], rs78322982[G], rs6499148[C], rs113717347[G], rs80356178[G], rs117400881[T], rs1477029[G], rs78831406[C], rs4784276[T], rs77419902[C], rs78724580[G], rs72812119[A], rs72812122[G], rs76365437[G], rs13335861[A], rs4352043[C], rs36099013[G], rs72812136[C], rs79815180[C], rs74018575[A], rs72812140[G], rs56085964[T], rs8051957[G], rs8050297[A], rs79721160[G], rs79088862[G], rs4784287[T], rs117297253[G], rs72797618[C], rs9933155[T], rs56278234[C], rs3931698[T], rs62049054[T], rs113465432[C], rs11641638[G] |

Continued on next page

**Supplementary Table S5**: Haplotype alleles (Continued)

| Variable | SNP[allele] |
|---|---|
| h3 | rs1420281[T], rs60526343[T], rs72792181[G], rs9939929[C], rs3095540[T], rs116988362[T], rs4785118[G], rs17268400[T], rs72792197[A], rs117285516[C], rs116048640[C], rs7205520[A], rs118147607[C], rs113755361[C], rs113083456[C], rs12930272[G], rs1345316[A], rs2287071[A], rs2540701[G], rs112797109[C], rs72794126[C], rs74393050[G], rs17270950[T], rs117142114[T], rs117832727[G], rs4784165[T], h1, rs79291013[C], rs117746278[G], rs79993873[A], rs78320298[A], rs62041381[A], rs117222080[G], rs117551222[T], rs7500472[G], rs72810045[G], rs12446016[T], rs73591253[C], rs150704739[C], rs113964117[G], rs142367768[C], rs117911537[C], rs16951771[G], rs73597288[G], rs12919591[A], rs6499147[C], rs78322982[G], rs62043282[C], rs113717347[G], rs62043323[G], rs80356178[G], rs34261756[T], rs117400881[T], rs4784276[T], rs77419902[C], rs78724580[G], rs72812119[A], rs72812122[G], rs76365437[G], rs13335861[A], rs4352043[C], rs36099013[G], rs72812136[C], rs74018575[A], rs72812140[G], rs34382563[C], rs12598778[T], rs79088862[G], rs4784287[T], rs117297253[G], rs56278234[C], rs3931698[T], rs113465432[C], rs11641638[G] |

Continued on next page

**Supplementary Table S5**: Haplotype alleles (Continued)

| Variable | SNP[allele] |
|---|---|
| h4 | rs60526343[T], rs72792181[G], rs9939929[C], rs116988362[T], rs4785118[G], rs17268400[T], rs72792197[A], rs117285516[C], rs116048640[C], rs7205520[A], rs118147607[C], rs113755361[C], rs113083456[C], rs12930272[G], rs2540701[G], rs112797109[C], rs72794126[C], rs74393050[G], rs17270950[T], rs117142114[T], rs117832727[G], rs4784165[T], h1, rs79291013[C], rs8044632[T], rs117746278[A], rs9939896[C], rs2024446[G], rs17298178[T], rs79993873[A], rs78320298[A], rs62041381[A], rs117222080[G], rs117551222[T], rs7500472[G], rs72810045[G], rs12446016[T], rs73591253[C], rs150704739[C], rs113964117[G], rs1345327[T], rs142367768[C], rs4783791[T], rs117911537[C], rs1344484[T], rs16951771[G], rs73597288[G], rs12919591[A], rs9630629[C], rs6499147[C], rs12922473[A], rs78322982[G], rs62043282[C], rs62043323[G], rs80356178[G], rs34261756[T], rs117400881[T], rs78831406[C], rs4784276[T], rs77419902[C], rs78724580[G], rs72812119[A], rs72812122[G], rs76365437[G], rs13335861[A], rs4352043[C], rs36099013[G], rs72812136[C], rs79815180[C], rs72812140[G], rs34382563[C], rs8050297[A], rs79721160[G], rs79088862[G], rs4784287[T], rs117297253[G], rs3931698[T], rs62049054[T], rs113465432[C], rs11641638[G] |

Continued on next page

**Supplementary Table S5**: Haplotype alleles (Continued)

| Variable | SNP[allele] |
|---|---|
| h5 | rs1420281[T], rs72792181[G], rs9939929[C], rs3095540[T], rs11649329[C], rs116988362[T], rs4785118[G], rs17268400[T], rs117108822[C], rs72792197[A], rs117285516[C], rs116048640[C], rs7205520[A], rs118147607[C], rs113755361[C], rs113083456[C], rs1345316[A], rs2540701[G], rs112797109[C], rs72794126[C], rs74393050[G], rs17270950[T], rs117142114[T], rs117832727[G], rs9938443[A], h1, rs79291013[C], rs117746278[G], rs17298178[T], rs79993873[A], rs78320298[A], rs62041381[A], rs117222080[G], rs7500472[G], rs72810045[G], rs12446016[T], rs73591253[C], rs150704739[C], rs113964117[G], rs142367768[C], rs4783791[T], rs1344484[T], rs73597288[G], rs12922473[A], rs78322982[G], rs62043282[C], rs113717347[G], rs62043323[G], rs80356178[G], rs34261756[T], rs117400881[T], rs1477029[G], rs78831406[C], rs4784276[T], rs77419902[T], rs78724580[G], rs72812119[A], rs72812122[G], rs62047145[C], rs76365437[G], rs13335861[A], rs4352043[C], rs36099013[G], rs79815180[C], rs34382563[C], rs12598778[T], rs12922187[A], rs8051957[A], rs79721160[G], rs79088862[G], rs4784287[T], rs117297253[G], rs3931698[T], rs62049054[T], rs113465432[C] |

Continued on next page

**Supplementary Table S5**: Haplotype alleles (Continued)

| Variable | SNP[allele] |
|---|---|
| h6 | rs1420281[T], rs60526343[T], rs72792181[G], rs9939929[C], rs3095540[T], rs11649329[T], rs116988362[T], rs4785118[G], rs17268400[T], rs117108822[C], rs72792197[A], rs117285516[C], rs116048640[C], rs118147607[C], rs113755361[C], rs1345316[A], rs2287071[A], rs112797109[C], rs72794126[C], rs74393050[G], rs17270950[T], rs117142114[T], rs117832727[G], rs9938443[A], rs4784165[G], h1, rs79291013[C], rs117746278[G], rs17298178[T], rs79993873[A], rs78320298[A], rs62041381[A], rs117222080[G], rs117551222[T], rs7500472[G], rs72810045[G], rs150704739[C], rs1345327[C], rs142367768[C], rs117911537[C], rs16951771[G], rs73597288[G], rs12919591[A], rs9630629[A], rs6499147[C], rs78322982[G], rs6499148[A], rs62043282[C], rs113717347[G], rs62043323[G], rs12924810[C], rs80356178[G], rs34261756[T], rs1477029[A], rs7199322[T], rs78831406[C], rs4784276[T], rs77419902[C], rs78724580[G], rs72812119[A], rs72812122[G], rs62047145[C], rs13335861[A], rs4352043[C], rs36099013[G], rs72812136[C], rs74018575[A], rs72812140[G], rs34382563[C], rs12598778[T], rs56085964[C], rs7499206[C], rs12922187[A], rs79721160[G], rs79088862[G], rs4784287[T], rs117297253[G], rs72797618[T], rs9933155[C], rs3931698[T], rs62049054[C] |

Continued on next page

**Supplementary Table S5**: Haplotype alleles (Continued)

| Variable | SNP[allele] |
|---|---|
| h7 | rs1420281[C], rs60526343[T], rs72792181[G], rs9939929[C], rs116988362[T], rs4785118[G], rs17268400[T], rs117108822[C], rs72792197[A], rs117285516[C], rs116048640[C], rs7205520[A], rs118147607[C], rs113755361[C], rs113083456[C], rs2287071[A], rs2540701[G], rs112797109[C], rs72794126[C], rs74393050[G], rs17270950[T], rs117142114[T], rs117832727[G], h1, rs79291013[C], rs117746278[G], rs9939896[C], rs79993873[A], rs78320298[A], rs62041381[A], rs117222080[G], rs117551222[T], rs7500472[G], rs12446016[T], rs73591253[C], rs150704739[C], rs113964117[G], rs1345327[C], rs117911537[C], rs16951771[G], rs73597288[G], rs12919591[A], rs9630629[A], rs78322982[G], rs113717347[G], rs62043323[G], rs12924810[C], rs80356178[G], rs117400881[T], rs7199322[T], rs78831406[C], rs4784276[T], rs77419902[C], rs78724580[G], rs72812119[A], rs72812122[G], rs62047145[C], rs76365437[G], rs13335861[A], rs4352043[C], rs36099013[G], rs72812136[C], rs79815180[C], rs74018575[A], rs7499206[C], rs12922187[A], rs8051957[G], rs8050297[A], rs79721160[G], rs79088862[G], rs4784287[T], rs9933155[T], rs56278234[C], rs3931698[T], rs62049054[T], rs113465432[C] |

Continued on next page

**Supplementary Table S5**: Haplotype alleles (Continued)

| Variable | SNP[allele] |
|---|---|
| h8 | rs1420281[T], rs60526343[T], rs72792181[G], rs9939929[C], rs3095540[T], rs11649329[T], rs4785118[G], rs17268400[T], rs117108822[C], rs72792197[A], rs117285516[C], rs116048640[C], rs118147607[C], rs113755361[C], rs113083456[C], rs1345316[A], rs2287071[A], rs112797109[C], rs72794126[C], rs74393050[G], rs17270950[T], rs117142114[T], rs117832727[G], h1, rs79291013[C], rs8044632[T], rs117746278[G], rs9939896[C], rs2024446[G], rs17298178[T], rs79993873[A], rs78320298[A], rs62041381[A], rs117222080[G], rs117551222[T], rs7500472[G], rs72810045[G], rs12446016[T], rs73591253[C], rs150704739[C], rs113964117[G], rs1345327[T], rs142367768[C], rs4783791[T], rs117911537[C], rs1344484[T], rs16951771[G], rs73597288[G], rs12919591[A], rs9630629[C], rs6499147[C], rs12922473[A], rs78322982[G], rs62043282[C], rs113717347[G], rs62043323[G], rs80356178[G], rs34261756[T], rs117400881[T], rs1477029[G], rs78831406[C], rs4784276[T], rs78724580[G], rs72812119[A], rs72812122[G], rs62047145[C], rs76365437[G], rs13335861[A], rs4352043[C], rs36099013[G], rs72812136[C], rs79815180[C], rs34382563[C], rs12598778[T], rs8050297[A], rs79088862[G], rs4784287[T], rs117297253[G], rs3931698[T], rs62049054[T] |

Continued on next page

**Supplementary Table S5**: Haplotype alleles (Continued)

| Variable | SNP[allele] |
|---|---|
| h9 | rs1420281[T], rs60526343[T], rs72792181[G], rs9939929[C], rs3095540[T], rs116988362[T], rs4785118[G], rs17268400[T], rs117108822[C], rs72792197[A], rs117285516[C], rs7205520[A], rs118147607[C], rs113755361[C], rs113083456[C], rs12930272[G], rs1345316[A], rs2287071[A], rs1362376[T], rs2540701[A], rs112797109[C], rs72794126[C], rs74393050[G], rs17270950[T], rs117142114[T], rs117832727[G], rs9938443[A], rs4784165[G], h1, rs79291013[C], rs8044632[C], rs117746278[G], rs9939896[T], rs2024446[A], rs17298178[T], rs79993873[A], rs78320298[A], rs62041381[A], rs117551222[T], rs7500472[G], rs72810045[G], rs73591253[C], rs150704739[C], rs113964117[G], rs142367768[C], rs117911537[C], rs16951771[G], rs73597288[G], rs12919591[A], rs6499147[C], rs78322982[G], rs62043282[C], rs113717347[G], rs62043323[G], rs12924810[C], rs80356178[G], rs34261756[T], rs117400881[T], rs7199322[T], rs78831406[C], rs4784276[T], rs78724580[G], rs72812119[A], rs72812122[G], rs76365437[G], rs13335861[A], rs4352043[C], rs36099013[G], rs72812136[C], rs79815180[C], rs72812140[G], rs34382563[C], rs12598778[T], rs56085964[T], rs7499206[T], rs79721160[G], rs79088862[G], rs117297253[G], rs72797618[T], rs9933155[C], rs56278234[T], rs3931698[T], rs62049054[T], rs113465432[C] |

Continued on next page

**Supplementary Table S5**: Haplotype alleles (Continued)

| Variable | SNP[allele] |
|---|---|
| h10 | rs1420281[T], rs72792181[G], rs9939929[C], rs11649329[C], rs116988362[T], rs4785118[G], rs17268400[T], rs117108822[C], rs72792197[A], rs117285516[C], rs116048640[C], rs7205520[A], rs118147607[C], rs113755361[C], rs113083456[C], rs2287071[A], rs2540701[G], rs112797109[C], rs72794126[C], rs74393050[G], rs17270950[T], rs117142114[T], rs117832727[G], rs4784165[T], h1, rs79291013[C], rs8044632[T], rs117746278[G], rs17298178[T], rs79993873[A], rs78320298[A], rs62041381[A], rs117222080[G], rs117551222[T], rs12446016[T], rs73591253[C], rs150704739[C], rs113964117[G], rs142367768[C], rs117911537[C], rs16951771[G], rs73597288[G], rs9630629[A], rs6499147[C], rs12922473[G], rs78322982[G], rs62043282[C], rs113717347[G], rs62043323[G], rs12924810[C], rs80356178[G], rs34261756[T], rs117400881[T], rs1477029[G], rs78831406[C], rs4784276[T], rs78724580[G], rs72812119[A], rs62047145[C], rs76365437[G], rs13335861[A], rs36099013[G], rs72812136[C], rs79815180[C], rs72812140[G], rs34382563[C], rs12598778[T], rs12922187[A], rs8050297[A], rs79721160[G], rs79088862[G], rs4784287[T], rs72797618[T], rs56278234[C], rs3931698[T], rs62049054[T], rs113465432[C], rs11641638[G] |

Continued on next page

**Supplementary Table S5**: Haplotype alleles (Continued)

| Variable | SNP[allele] |
|---|---|
| h11 | rs1420281[T], rs60526343[T], rs72792181[G], rs9939929[C], rs11649329[C], rs116988362[T], rs4785118[G], rs17268400[T], rs72792197[A], rs117285516[C], rs116048640[C], rs7205520[A], rs118147607[C], rs113755361[C], rs113083456[C], rs12930272[G], rs1345316[A], rs1362376[T], rs2540701[G], rs112797109[C], rs72794126[C], rs74393050[G], rs17270950[T], rs117142114[T], rs117832727[G], rs4784165[T], h1, rs79291013[C], rs8044632[C], rs117746278[G], rs9939896[T], rs2024446[A], rs17298178[T], rs79993873[A], rs78320298[A], rs62041381[A], rs117222080[G], rs117551222[T], rs72810045[G], rs12446016[T], rs73591253[C], rs150704739[C], rs113964117[G], rs1345327[C], rs142367768[C], rs4783791[G], rs117911537[C], rs1344484[C], rs16951771[G], rs73597288[G], rs12919591[A], rs9630629[A], rs6499147[C], rs12922473[G], rs78322982[G], rs6499148[A], rs62043282[C], rs113717347[G], rs62043323[G], rs12924810[C], rs80356178[G], rs34261756[T], rs117400881[T], rs7199322[T], rs78831406[C], rs4784276[T], rs78724580[G], rs72812119[A], rs72812122[G], rs62047145[C], rs76365437[G], rs13335861[A], rs4352043[C], rs36099013[G], rs79815180[C], rs34382563[C], rs12598778[T], rs12922187[A], rs8051957[A], rs79721160[G], rs79088862[G], rs4784287[T], rs117297253[G], rs72797618[T], rs9933155[C], rs3931698[T], rs62049054[C], rs11641638[G] |

Continued on next page

**Supplementary Table S5**: Haplotype alleles (Continued)

| Variable | SNP[allele] |
|---|---|
| h12 | rs1420281[T], rs60526343[T], rs72792181[G], rs9939929[C], rs3095540[T], rs11649329[T], rs116988362[T], rs4785118[G], rs17268400[T], rs117108822[C], rs72792197[A], rs117285516[C], rs116048640[C], rs118147607[C], rs113755361[C], rs113083456[C], rs12930272[G], rs1345316[A], rs1362376[T], rs112797109[C], rs17270950[T], rs117142114[T], rs117832727[G], rs9938443[A], rs4784165[T], h1, rs79291013[C], rs117746278[G], rs17298178[T], rs79993873[A], rs78320298[A], rs62041381[A], rs117222080[G], rs117551222[T], rs7500472[G], rs72810045[G], rs12446016[T], rs150704739[C], rs1345327[C], rs142367768[C], rs117911537[C], rs16951771[G], rs73597288[G], rs12919591[A], rs9630629[A], rs6499147[C], rs12922473[G], rs78322982[G], rs6499148[C], rs113717347[G], rs62043323[G], rs80356178[G], rs117400881[T], rs1477029[G], rs78831406[C], rs4784276[T], rs77419902[C], rs78724580[G], rs72812119[A], rs72812122[G], rs76365437[G], rs13335861[A], rs4352043[C], rs36099013[G], rs72812136[C], rs79815180[C], rs72812140[G], rs56085964[T], rs12922187[A], rs8050297[A], rs79721160[G], rs79088862[G], rs4784287[T], rs117297253[G], rs72797618[T], rs3931698[T], rs62049054[T], rs113465432[C], rs11641638[G] |

Supplementary Table S6 gives the frequencies of the discovered haplotypes in controls in UKBB and DRIVE for the purposes of combining controls during replication. Frequencies were compared using Fisher's Exact Test. Haplotypes with $P$-values less than 0.05 were considered significantly different and were not combined across populations.

**Supplementary Table S6**: Controls frequency differences

| Variable | UKBB relative frequency[a] $/10^{-4}$ | DRIVE relative frequency[b] $/10^{-4}$ | Fisher $P$-value |
|---|---|---|---|
| | Chr10 | | |
| h2 | 0.2616 | 1.582 | $6.0 \times 10^{-4}$ |
| h3 | 0.5831 | 0.1978 | 0.51 |
| h4 | 4.825 | 4.549 | 0.91 |
| h5 | 0.6685 | 0.7911 | 0.77 |
| h6 | 1.105 | 1.780 | 0.19 |
| h7 | 0.4651 | 0.1978 | 0.71 |
| | Chr11 | | |
| h2 | 3.372 | 4.351 | 0.25 |
| h3 | 1.424 | 4.351 | $5.0 \times 10^{-5}$ |
| h4 | 0.1453 | 0.1978 | 0.56 |
| | Chr16 | | |
| h2 | 0.0 | 0.0 | 1.0 |
| h3 | 1.773 | 1.384 | 0.72 |
| h4 | 0.8720 | 1.582 | 0.14 |
| h5 | 7.092 | 7.713 | 0.59 |
| h6 | 0.1163 | 0.1978 | 0.50 |
| h7 | 1.105 | 2.175 | 0.053 |
| h8 | 9.621 | 5.933 | $9.1 \times 10^{-3}$ |
| h9 | 0.1453 | 0.1978 | 0.56 |
| h10 | 0.02907 | 0.1978 | 0.24 |
| h11 | 1.453 | 0.7911 | 0.31 |
| h12 | 0.8720 | 1.780 | 0.087 |

[a] Among 344,046 controls chromosomes

[b] Among 50,564 controls chromosomes

#### 6. Principal components analysis

All statistical models were adjusted for the first ten genotype principal components. PCA on all UKBB samples was performed by the UK Biobank on 407,219 individuals at 147,604 genotyped markers [1]; the first three PCs of the 181,034 "white British" females used in the discovery analysis are plotted in Supplementary Fig. S2. Next, PCA was performed using FlashPCA [2] on 59,958 DRIVE women together 1000 Genomes Phase 3 individuals [3] at 193,786 LD-pruned markers to identify 55,450 unrelated DRIVE women of European ancestry (Supplementary Fig. S3). The DRIVE European cluster was defined by the cutoffs PC1 $\leq 0.0025$ and PC2 $\geq -0.007$. Finally, genotype principal components used in the replication analyses were generated for 55,346 DRIVE subjects and the combined 181,034 + 55,346 = 236,080 UKBB and DRIVE subjects, respectively, at 101,013 and 1,243,728 LD-pruned markers (Supplementary Figs. S4 and S5). Note that the magnitudes of the PCs for the UKBB-provided data are approximately 100-fold larger than the ones computed in this study.

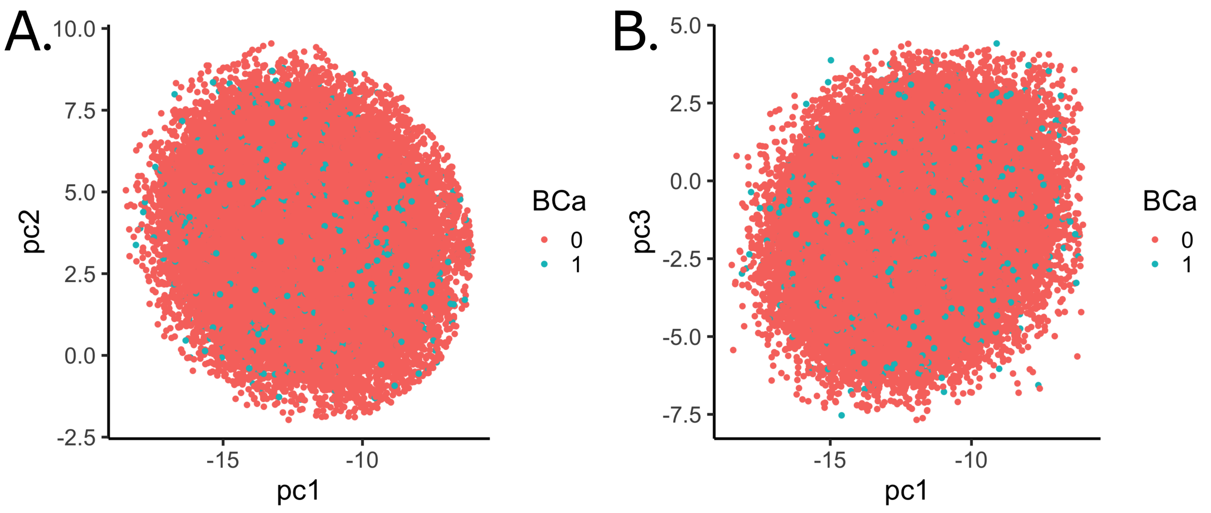

**Supplementary Figure S2:** Plots of PC2 vs. PC1 (A) and PC3 vs. PC1 (B) for 181,034 UKBB subjects, stratified by breast cancer (BCa) status, computed using 147,604 genotyped markers.

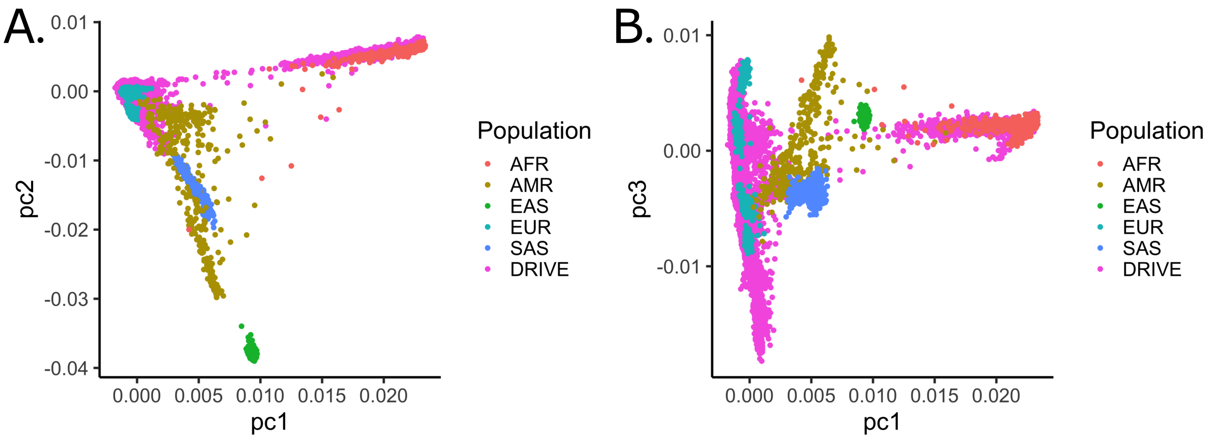

**Supplementary Figure S3:** Plots of PC2 vs. PC1 (A) and PC3 vs. PC1 (B) for 62,462 DRIVE subjects and 1000 Genomes Phase 3 individuals, stratified by ancestry, computed using 193,786 LD-pruned markers.

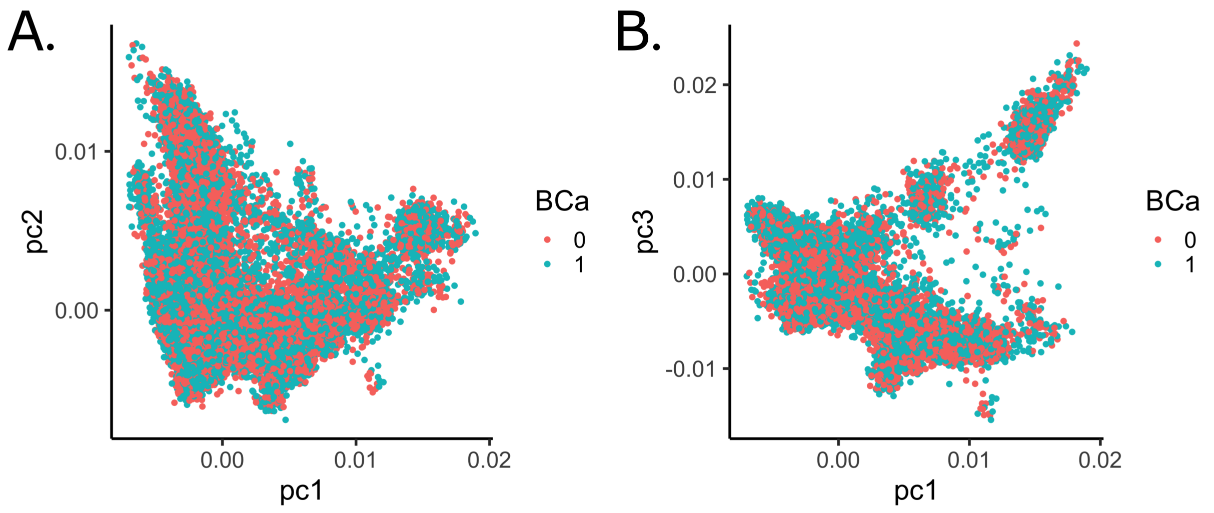

**Supplementary Figure S4:** Plots of PC2 vs. PC1 (A) and PC3 vs. PC1 (B) for 55,346 DRIVE subjects, stratified by breast cancer (BCa) status, computed using 101,013 LD-pruned markers.

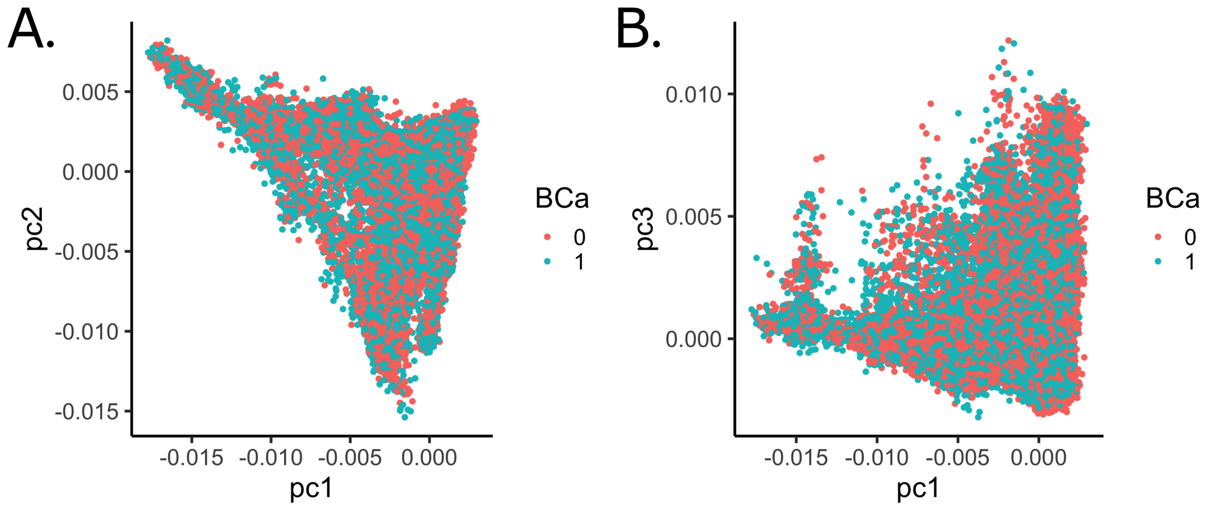

**Supplementary Figure S5:** Plots of PC2 vs. PC1 (A) and PC3 vs. PC1 (B) for 236,380 UKBB and DRIVE subjects, stratified by breast cancer (BCa) status, computed using 1,243,728 LD-pruned markers.

The coefficients of the genotype principal components in the null model of each of the discovery and replication analyses are given in Supplementary Table S7. Age was taken as a covariate in the replication analyses where logistic regression was used.

**Supplementary Table S7**: Effect sizes of the genotype principal components in the baseline models

| Variable | HR/OR[a] (95% CI) | $P$-value[b] |
|---|---|---|
| | UKBB discovery | |
| pc1 | 0.999 (0.986–1.01) | 0.88 |
| pc2 | 0.986 (0.972–1.00) | 0.045 |
| pc3 | 1.01 (0.994–1.02) | 0.30 |
| pc4 | 1.00 (0.993–1.01) | 0.50 |
| pc5 | 1.00 (0.995–1.00) | 0.82 |
| pc6 | 1.01 (0.994–1.02) | 0.29 |
| pc7 | 1.01 (0.994–1.02) | 0.35 |
| pc8 | 0.999 (0.988–1.01) | 0.86 |
| pc9 | 0.998 (0.993–1.00) | 0.28 |
| pc10 | 1.01 (0.996–1.02) | 0.26 |
| | DRIVE replication | |
| age | 1.02 (1.02–1.02) | $5.1 \times 10^{-114}$ |
| pc1 | 1.80 (0.0334–96.9) | 0.77 |
| pc2 | $3.62 \times 10^{-6}$ ($6.89 \times 10^{-8}$–$1.91 \times 10^{-4}$) | $5.8 \times 10^{-10}$ |
| pc3 | $1.88 \times 10^{3}$ (32.6–$1.08 \times 10^{5}$) | $2.6 \times 10^{-4}$ |
| pc4 | $1.33 \times 10^{4}$ (252–$7.01 \times 10^{5}$) | $2.7 \times 10^{-6}$ |
| pc5 | $3.59 \times 10^{-3}$ ($6.71 \times 10^{-5}$–0.19) | $5.6 \times 10^{-3}$ |
| pc6 | 0.20 ($3.67 \times 10^{-3}$–10.6) | 0.42 |
| pc7 | $2.16 \times 10^{4}$ (409–$1.14 \times 10^{6}$) | $8.2 \times 10^{-7}$ |
| pc8 | $1.58 \times 10^{-3}$ ($2.98 \times 10^{-5}$–0.0837) | $1.4 \times 10^{-3}$ |
| pc9 | $8.10 \times 10^{-14}$ ($1.45 \times 10^{-15}$–$4.51 \times 10^{-12}$) | $6.8 \times 10^{-49}$ |
| pc10 | 0.0396 ($7.44 \times 10^{-4}$–2.11) | 0.11 |
| | Combined DRIVE+UKBB replication | |
| age | 0.987 (0.986 – 0.988) | $2.0 \times 10^{-70}$ |
| pc1 | $7.36 \times 10^{-154}$ ($3.67 \times 10^{-156}$–$1.47 \times 10^{-151}$) | 0.0 |
| pc2 | $2.80 \times 10^{-105}$ ($1.56 \times 10^{-107}$–$5.03 \times 10^{-103}$) | 0.0 |
| pc3 | $2.91 \times 10^{11}$ ($2.79 \times 10^{8}$–$3.05 \times 10^{14}$) | $9.9 \times 10^{-14}$ |
| pc4 | $9.98 \times 10^{-34}$ ($9.93 \times 10^{-37}$–$1.00 \times 10^{-30}$) | $6.1 \times 10^{-103}$ |
| pc5 | $9.21 \times 10^{-36}$ ($7.97 \times 10^{-39}$–$1.06 \times 10^{-32}$) | $2.5 \times 10^{-111}$ |
| pc6 | $1.09 \times 10^{6}$ ($1.26 \times 10^{3}$–$9.43 \times 10^{8}$) | $5.6 \times 10^{-5}$ |

Continued on next page

**Supplementary Table S7**: Effect sizes of the genotype principal components in the baseline models (Continued)

| Variable | HR/OR[a] (95% CI) | $P$-value[b] |
| --- | --- | --- |
| pc7 | $8.14 \times 10^{-5}$ ($1.06 \times 10^{-7}$–0.062) | $5.5 \times 10^{-3}$ |
| pc8 | $4.27 \times 10^{6}$ ($6.86 \times 10^{3}$–$2.66 \times 10^{9}$) | $3.3 \times 10^{-6}$ |
| pc9 | $2.22 \times 10^{-15}$ ($2.87 \times 10^{-18}$–$1.71 \times 10^{-12}$) | $2.6 \times 10^{-23}$ |
| pc10 | $1.22 \times 10^{12}$ ($1.09 \times 10^{9}$–$1.36 \times 10^{15}$) | $7.5 \times 10^{-15}$ |

[a] HR in UKBB discovery, OR in DRIVE and DRIVE+UKBB replication

[b] Wald test $P$-value

#### 7. LD blocks

LD blocks based on the 1000 Genomes Phase 3 [3] EUR population were estimated using Big-LD [4]. Pruning parameters were `MAF` = 0.05 and `appendRare` = `FALSE`. PLINK input files (ped and map) were generated for the regions 10:120850027–121949982 (32,464 SNPs, 2,524 with `MAF` < 0.05), 11:69050018–69699989 (21,238 SNPs, 1,470 with `MAF` < 0.05), and 16:52000011–53099995 (33,731 SNPs, 2,308 with `MAF` < 0.05). Results based on the $r^2$ metric are shown in Supplementary Figs. S6–S8.

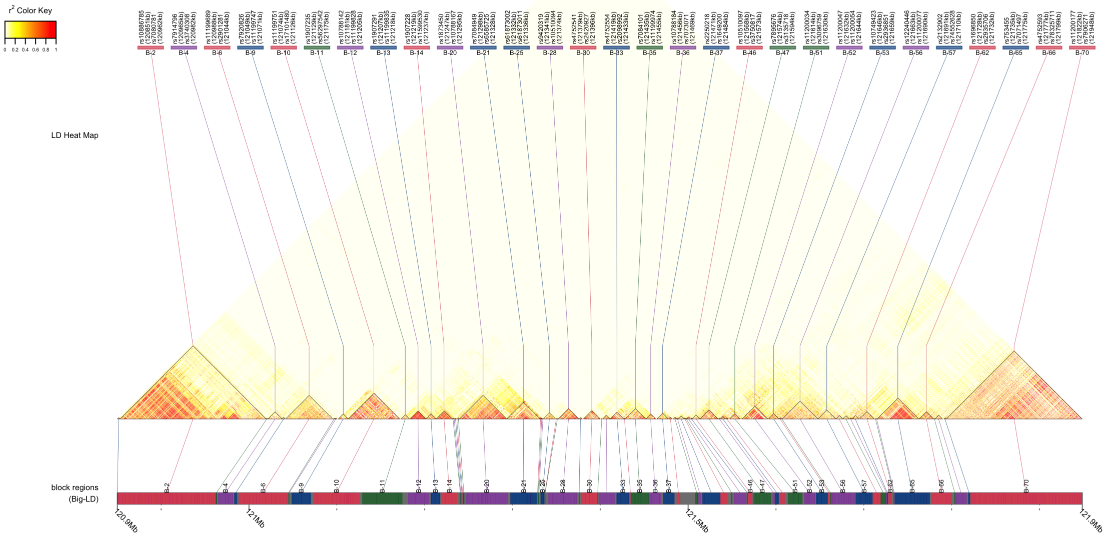

**Supplementary Figure S6:** 1000 Genomes Phase 3 EUR LD blocks at chromosome locus 10q26, spanning 2,524 SNPs in 10:120850027–121949982.

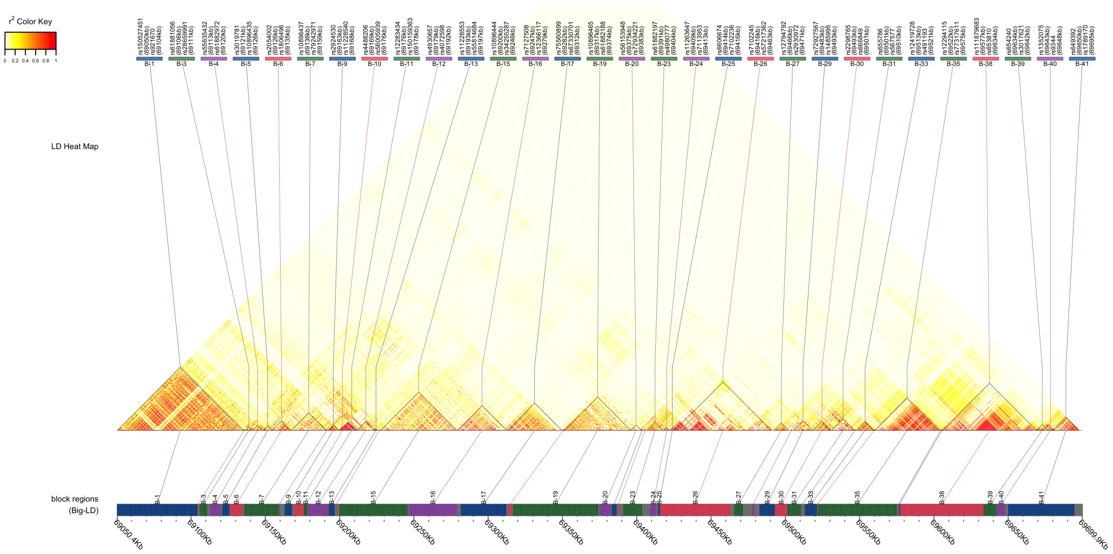

**Supplementary Figure S7:** 1000 Genomes Phase 3 EUR LD blocks at chromosome locus 11q13, spanning 1,470 SNPs in 11:69050018–69699989.

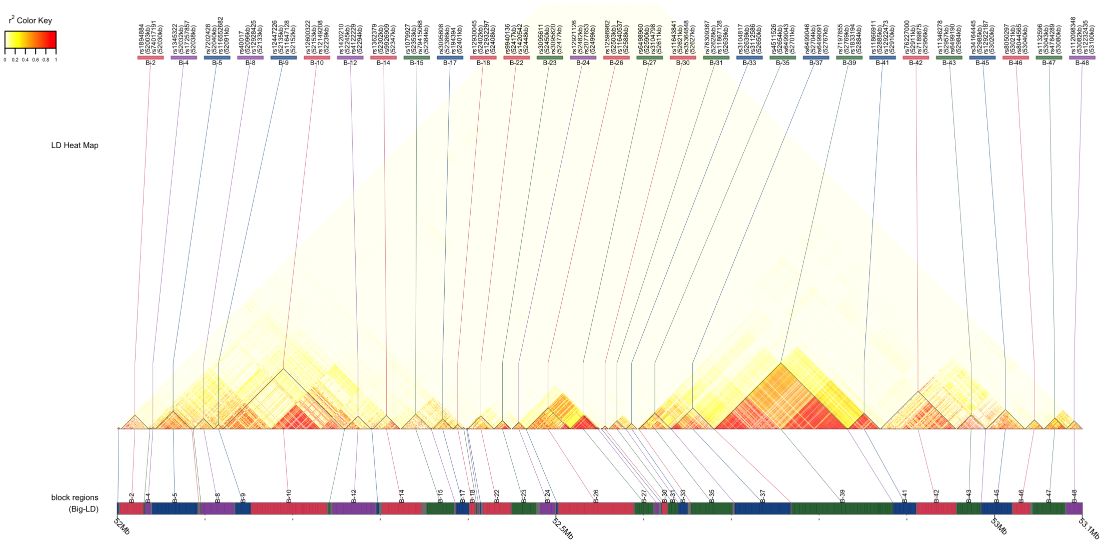

**Supplementary Figure S8:** 1000 Genomes Phase 3 EUR LD blocks at chromosome locus 16q12, spanning 2,308 SNPs in 16:52000011–53099995.

To determine whether the rare haplotypes could have been due to rare recombination events, we measured the frequency of each segment of the replicated haplotypes by LD block. Specifically, we measured the frequency of the subset of the rare haplotype lying in each LD block identified by Big-LD in UKBB. The results are in Supplementary Table S8.

**Supplementary Table S8**: Haplotype segment frequencies by LD blocks

| Range | Segment | UKBB frequency |
|---|---|---|
| | Chr10 | |
| h5 | | |
| chr10:121048540–121070530 | rs4752498[T] | 0.852475 |
| chr10:121073631–121125637 | rs7078330[C], rs76468001[C], rs117641945[T], rs78493319[C], rs118104738[C], rs2130789[T], rs72826685[T], rs10886849[C], rs2244506[T], rs185196870[C], rs7907754[A], rs72828403[A], rs2254069[G] | 0.56944 |
| chr10:121129041–121174596 | rs78250596[G], rs77170423[T], rs12259125[G], rs12777051[C], rs17101493[T], rs2901286[C], rs80274247[T], rs61874880[G], rs11199800[A], rs11199805[C], rs71484661[C], rs56297542[C] | 0.224154 |
| chr10:121181315–121205116 | rs11597363[C], rs11199825[G] | 0.885635 |
| chr10:121206945-121217643 | rs12777794[C], rs12221275[C], rs76847969[T], rs75273759[C], rs80351942[G], rs1907227[T] | 0.738469 |
| chr10:121219430–121236985 | rs55740531[T], rs76105384[A], rs10886873[T] | 0.521617 |
| chr10:121244014–121245125 | rs116967076[G], rs7899480[C] | 0.731967 |
| chr10:121246760–121294504 | rs11199860[A], rs7076500[A], rs4752526[T], rs12253779[A], rs78331385[C], rs80091641[C], rs77914714[T] | 0.37161 |
| chr10:121297678–121327951 | rs12570783[A], rs75707486[G], rs117666755[G] | 0.548773 |
| chr10:121328885–121329695 | rs76866916[G] | 0.988071 |
| chr10:121332029–121336400 | rs4752536[A], rs116988595[T], rs35098964[C] | 0.32991 |
| chr10:121337205–121337233 | rs10466215[T] | 0.748757 |
| chr10:121340748–121373689 | rs146809981[A], rs7893305[A], rs12256422[G], rs80126005[G], rs7071430[C], rs2420932[T], rs60348293[G], rs17601696[C], rs12220114[G] | 0.0883066 |
| chr10:121375686–121376246 | rs10788173[C] | 0.386662 |

Continued on next page

**Supplementary Table S8**: Haplotype segment frequencies by LD blocks (Continued)

| Range | Segment | UKBB frequency |
|---|---|---|
| chr10:121378743–121396482 | rs1896403[A], rs73362352[G], rs76834389[C], rs4752550[C] | 0.541998 |
| chr10:121398221–121416615 | rs74158584[A], rs2162541[C], rs12413779[T], rs74158588[C], rs76352137[C] | 0.919076 |
| chr10:121418512–121432971 | rs11199954[G], rs17603569[A], rs11199957[C], rs17101979[T] | 0.841585 |
| chr10:121434209–121434484 | rs11199964[T] | 0.730388 |
| chr10:121487098–121490734 | rs3135795[G] | 0.992921 |
| chr10:121508117–121511625 | rs116941692[C] | 0.984287 |
| chr10:121552184–121566241 | rs56226109[G] | 0.995512 |
| chr10:121574483–121594355[a] | rs2981579[A], rs45631549[C], rs45631614[G], rs45631630[G], rs17542858[G] | 0.384312 |
| chr10:121645507–121659103 | rs118181559[G] | 0.9594 |
| | Chr11 | |
| h2 | | |
| chr11:69050429–69104171 | rs2376558[C], rs149949063[A], rs144905179[G], rs74674490[A], rs72930627[C], rs72930631[T] | 0.753386 |
| chr11:69105841–69110843 | rs11228502[T] | 0.730893 |
| chr11:69112649–69119685 | rs12577213[C], rs73520512[C], rs10896431[C] | 0.752461 |
| chr11:69126293–69134803 | rs74897859[C], rs55703863[G] | 0.838884 |
| chr11:69135997–69159087 | rs17149775[A], rs117236867[A], rs11228530[A], rs17309046[T], rs56134379[G], rs72932540[A] | 0.539644 |
| chr11:69163017–69168084 | rs78033785[C] | 0.901309 |
| chr11:69169026–69175864 | rs74342245[G] | 0.903386 |
| chr11:69178893–69192221 | rs35435383[G] | 0.954616 |
| chr11:69192716–69196670 | rs117694794[C] | 0.979015 |
| chr11:69199780–69245668 | rs78208050[A], rs61881109[G], rs75590802[G], rs7937094[C], rs7931342[T], rs10896449[A], rs7130881[A] | 0.431402 |
| chr11:69246550–69279008 | rs7119988[G], rs111762835[G], rs4930672[G], rs11228599[G], rs7940107[G], rs72930267[A], rs7946255[T], rs116951063[C], rs118004919[G], rs35637432[G], rs34737133[C], rs11825015[C] | 0.653913 |

Continued on next page

**Supplementary Table S8**: Haplotype segment frequencies by LD blocks (Continued)

| Range | Segment | UKBB frequency |
|---|---|---|
| chr11:69281620–69312240 | rs78656034[C], rs117821797[C], rs56103266[G], rs117131950[G], rs7103126[C], rs116926312[C], rs12274095[C], rs10896461[G], rs78919175[C], rs7124547[T], rs12789955[T] | 0.0859093 |
| chr11:69316676–69373986 | rs115223540[C], rs28378931[A], rs117186144[C], rs72932198[G], rs12802601[T], rs12796465[A] | 0.832346 |
| chr11:69374868–69382544 | rs61882193[A] | 0.836661 |
| chr11:69383219–69386345 | rs11263638[G] | 0.941196 |
| chr11:69414366–69415653 | rs74471298[G] | 0.963728 |
| chr11:69415686–69463122 | rs7105934[G], rs79373485[C], rs4444099[C] | 0.869605 |
| chr11:69482794–69492939 | rs1122316[G] | 0.842066 |
| chr11:69501433–69510343 | rs76809977[C] | 0.901532 |
| chr11:69513149–69521223[a] | rs559664[G], rs657315[C] | 0.120499 |
| chr11:69521958–69574960 | rs498931[A], rs71465432[C], rs183782062[G], rs79442425[C], rs111929748[G], rs117490805[G] | 0.761995 |
| chr11:69577175–69633526 | rs79241527[C], rs72932500[A], rs57162717[A] | 0.930071 |
| h4 | | |
| chr11:69050429–69104171 | rs2376558[C], rs61881030[C], rs149949063[A], rs144905179[G], rs74674490[A] | 0.88777 |
| chr11:69112649–69119685 | rs12577213[C], rs73520512[C], rs3892895[G] | 0.668825 |
| chr11:69120988–69125890 | rs7116054[C] | 0.80365 |
| chr11:69126293–69134803 | rs74897859[C], rs17308715[C], rs67039008[A], rs55703863[G] | 0.445499 |
| chr11:69135997–69159087 | rs117236867[A], rs11228530[A], rs17309046[T], rs56134379[G] | 0.780166 |
| chr11:69163017–69168084 | rs78033785[C] | 0.901309 |
| chr11:69169026–69175864 | rs74342245[G] | 0.903386 |
| chr11:69178893–69192221 | rs35435383[G] | 0.954616 |
| chr11:69192716–69196670 | rs117694794[C] | 0.979015 |
| chr11:69199780–69245668 | rs78208050[A], rs61881109[G], rs75590802[G], rs11228565[G], rs7937094[C], rs7130881[A] | 0.678096 |

Continued on next page

**Supplementary Table S8**: Haplotype segment frequencies by LD blocks (Continued)

| Range | Segment | UKBB frequency |
|---|---|---|
| chr11:69246550–69279008 | rs7119988[G], rs111762835[G], rs4930672[G], rs11228599[G], rs7940107[G], rs72930267[A], rs7946255[T], rs116951063[C], rs118004919[G], rs35637432[G], rs34737133[C], rs11825015[C] | 0.653913 |
| chr11:69281620–69312240 | rs78656034[C], rs74551015[G], rs117821797[C], rs12275849[T], rs56103266[G], rs117131950[G], rs116926312[C], rs11539762[G], rs142581206[G], rs11228610[G], rs12274095[C], rs12224376[G], rs7124547[T], rs11603814[G] | 0.109283 |
| chr11:69316676–69373986 | rs115223540[C], rs28378931[A], rs117186144[C], rs79487139[C], rs11600497[C], rs72932198[G], rs12802601[T], rs12796465[A], rs4275647[C] | 0.4643 |
| chr11:69374868–69382544 | rs61882193[A] | 0.836661 |
| chr11:69383219–69386345 | rs11263638[G] | 0.941196 |
| chr11:69390979–69403510 | rs11263641[T] | 0.803929 |
| chr11:69409116–69413146 | rs76855139[C] | 0.90467 |
| chr11:69414366–69415653 | rs74471298[G] | 0.963728 |
| chr11:69415686–69463122 | rs7105934[G], rs79373485[C], rs4444099[C] | 0.869605 |
| chr11:69482794–69492939 | rs1122316[G] | 0.842066 |
| chr11:69501433–69510343 | rs76809977[C] | 0.901532 |
| chr11:69513149–69521223[a] | rs559664[G], rs657315[C] | 0.120499 |
| chr11:69521958–69574960 | rs498931[A], rs71465432[C], rs183782062[G], rs79442425[C], rs111929748[G], rs117490805[G] | 0.761995 |
| chr11:69577175–69633526 | rs79241527[C], rs72932500[A], rs57162717[A] | 0.930071 |
| | Chr16 | |
| h6 | | |
| chr16:52152613–52238530 | rs1420281[T], rs60526343[T], rs72792181[G], rs9939929[C], rs3095540[T], rs11649329[T], rs116988362[T], rs4785118[G], rs17268400[T], rs117108822[C], rs72792197[A], rs117285516[C], rs116048640[C], rs118147607[C], rs113755361[C] | 0.236005 |
| chr16:52245207–52294090 | rs1345316[A], rs2287071[A], rs112797109[C], rs72794126[C], rs74393050[G], rs17270950[T] | 0.611385 |

Continued on next page

**Supplementary Table S8**: Haplotype segment frequencies by LD blocks (Continued)

| Range | Segment | UKBB frequency |
|---|---|---|
| chr16:52301860–52346529 | rs117832727[G], rs9938443[A], rs4784165[G] | 0.242924 |
| chr16:52482063–52498590 | rs78338509[A], rs12447430[T], rs45577538[G] | 0.928375 |
| chr16:52502973–52588200[a] | rs45542333[G], rs78268044[C], rs45454402[A], rs45512493[A], rs34750829[G], rs9931232[A], rs4594251[C], rs4784227[T] | 0.234771 |
| chr16:52589725–52610983 | rs45584434[G], rs45560737[C], rs45575339[C], rs45587544[G] | 0.901234 |
| chr16:52620651–52626825 | rs78841172[T] | 0.971594 |
| chr16:52627575–52638704 | rs111925335[A], rs75127968[G] | 0.960121 |
| chr16:52768849–52884010 | rs79291013[C], rs117746278[G], rs17298178[T], rs79993873[A], rs78320298[A], rs62041381[A], rs117222080[G], rs117551222[T], rs7500472[G], rs72810045[G], rs150704739[C], rs1345327[C], rs142367768[C], rs117911537[C] | 0.327052 |
| chr16:52884723–52910315 | rs73597288[G], rs12919591[A], rs9630629[A], rs6499147[C] | 0.415988 |
| chr16:52911464–52955880 | rs78322982[G], rs6499148[A], rs62043282[C], rs113717347[G], rs62043323[G], rs12924810[C], rs80356178[G], rs34261756[T], rs1477029[A], rs7199322[T], rs78831406[C], rs4784276[T] | 0.339677 |
| chr16:52956954–52984166 | rs77419902[C], rs78724580[G], rs72812119[A], rs72812122[G], rs62047145[C], rs13335861[A], rs4352043[C] | 0.550941 |
| chr16:52985049–53019595 | rs36099013[G], rs72812136[C], rs74018575[A], rs72812140[G], rs34382563[C], rs12598778[T], rs56085964[C], rs7499206[C], rs12922187[A] | 0.405788 |
| chr16:53020579–53040078 | rs79721160[G], rs79088862[G], rs4784287[T], rs117297253[G], rs72797618[T], rs9933155[C], rs3931698[T] | 0.339671 |

[a] Region contains GWAS hit

*Note*: SNPs may not appear in table if they fall between predicted LD-blocks

### 8. Power analysis

We estimate the power $1-\beta$ to detect a given haplotype OR by two methods based on Fisher's Exact Test. Let the observed haplotype frequency in population $i$ be $f_i$, the number of chromosomes with the haplotype divided by the total number $2n_i$ of chromosomes in population $i$. The fraction of subjects with at least one copy of the haplotype is $p_i = f_i^2 + 2f_i(1-f_i) \approx 2f_i$ when $f_i \ll 1$. Then the $2 \times 2$ table for exposure to the haplotype is

| | Cases | Controls | |
|---|---|---|---|
| haplotype | $p_1 n_1$ | $p_2 n_2$ | $x$ |
| no haplotype | $(1-p_1)n_1$ | $(1-p_2)n_2$ | $y$ |
| | $n_1$ | $n_2$ | $n = n_1 + n_2$ |

(2)

The observed OR for this table is $\frac{p_1(1-p_2)}{(1-p_1)p_2}$. From an observed OR and given controls frequency the required cases frequency is

$$p_1 = \frac{\mathrm{OR} \cdot p_2}{1 - p_2 + \mathrm{OR} \cdot p_2}.$$

Let $z = \frac{\log(\mathrm{OR})}{\mathrm{se}(\log(\mathrm{OR}))}$ be a standard normal variable. The null hypothesis $H_0 : z = 0$ is tested against the alternative $H_1 : z \neq 0$. First, for a given type I error rate $\alpha$, we calculate the critical value $x^*$ for the hypothesis $z > 0$ as

$$\begin{aligned} Z_{1-\alpha/2} - Z_\beta &= \frac{x^* - 0}{\mathrm{se}(\log(\mathrm{OR}))} - \frac{x^* - \log(\mathrm{OR})}{\mathrm{se}(\log(\mathrm{OR}))} \\ Z_\beta &= -\frac{\log(\mathrm{OR})}{\mathrm{se}(\log(\mathrm{OR}))} + Z_{1-\alpha/2}, \end{aligned} \tag{3}$$

where $Z_X$ is the $100X^{\mathrm{th}}$ percentile of the standard normal distribution and $\mathrm{se}(\log(\mathrm{OR})) = \sqrt{\frac{1}{n_1 p_1 (1-p_1)} + \frac{1}{n_2 p_2 (1-p_2)}}$ is the standard error of $z$. When $z < 0$ we similarly calculate

$$Z_{1-\beta} = -\frac{\log(\mathrm{OR})}{\mathrm{se}(\log(\mathrm{OR}))} + Z_{\alpha/2}. \tag{4}$$

The total power is calculated by inverting Eqs. (3) and (4) and summing the two values of $1-\beta$.

The previous method relies on the normality of the log-OR. To get the exact power we use the simulation method `power.fisher.test` implemented in R. To test the hypothesis $H_1 : p_1 \neq p_2$ against the null hypothesis $H_0 : p_1 = p_2$, simulate the table (2) by sampling from two binomial distributions with parameters $n_1, p_1$ and $n_2, p_2$. The power to detect $p_1 > p_2$ is the fraction of tables with $\frac{\log(\mathrm{OR})}{\mathrm{se}(\log(\mathrm{OR}))} > Z_{1-\alpha/2}$; the power to detect $p_1 < p_2$ is the fraction of tables with $\frac{\log(\mathrm{OR})}{\mathrm{se}(\log(\mathrm{OR}))} < Z_{\alpha/2}$; the total power is the sum. We simulate `nsim` $= 10,000$ tables for each test. Because the haplotypes we evaluate are so rare, the simulated exact method was the primary mode of assessing power. The results for seven at least marginally replicated haplotypes are shown in Supplementary Table S9.

**Supplementary Table S9**: Power to detect replicated haplotypes

| Variable | OR[a] | Controls relative frequency[a]/$10^{-4}$ | UKBB discovery power[b] | DRIVE replication power[c] | Combined replication power[d] |
|---|---|---|---|---|---|
| | | | Chr10 | | |
| h5 | 2.69 | 0.7911 | $4.5 \times 10^{-3}$ ($4.6 \times 10^{-3}$) | 0.40 (0.41) | 0.76 (0.85) |
| | | | Chr11 | | |
| h2 | 2.13 | 4.351 | 0.059 (0.068) | 0.87 (0.85) | 0.99 (1.0) |
| h4 | 11.9 | 0.1978 | 0.17 (0.34) | 0.93 (0.67) | 1.0 (1.0) |
| | | | Chr16 | | |
| h6 | 2.89 | 0.1978 | $3.0 \times 10^{-4}$ ($3.2 \times 10^{-4}$) | 0.059 (0.15) | 0.34 (0.37) |

[a] Observed in DRIVE

[b] Based on $n_1 = 9,011$ cases and $n_2 = 172,023$ controls; $\alpha = 1.0 \times 10^{-5}$

[c] Based on $n_1 = 30,064$ cases and $n_2 = 25,282$ controls; $\alpha = 0.05$

[d] Based on $n_1 = 30,064$ cases and $n_2 = 25,282 + 172,023 = 197,305$ controls; $\alpha = 0.05$

*Note*: Each cell expressed as "simulated exacted power (approximate power)"
